# A dose-finding study to guide use of verapamil as an adjunctive therapy in tuberculosis

**DOI:** 10.1101/2023.07.28.23293316

**Authors:** Chandrasekaran Padmapriyadarsini, John D. Szumowski, Nabila Akbar, Prema Shanmugasundaram, Anilkumar Jain, Marasamy Bathragiri, Manoranjan Pattnaik, Jyotirmayee Turuk, Ramesh Karunaianantham, Senthilkumar Balakrishnan, Sangamitra Pati, Hemanth K. Agibothu Kupparam, Manoj Kumar Rathore, Jegadeesh Raja, K. Raghu Naidu, John Horn, Laura Whitworth, Roger Sewell, Lalita Ramakrishnan, Soumya Swaminathan, Paul H. Edelstein

## Abstract

Induction of mycobacterial efflux pumps is a cause of *Mycobacterium tuberculosis* (Mtb) drug tolerance, a barrier to shortening antitubercular treatment. Verapamil inhibits Mtb efflux pumps that mediate tolerance to rifampin, a cornerstone of tuberculosis treatment. Verapamil’s mycobacterial efflux pump inhibition also limits Mtb growth in macrophages in the absence of antibiotic treatment. These findings suggest that verapamil could be used as an adjunctive therapy for TB treatment shortening. However, verapamil is rapidly and substantially metabolized when co-administered with rifampin. We determined in a dose-escalation clinical trial that rifampin-induced clearance of verapamil can be countered without toxicity by the administration of larger than usual doses of verapamil. An oral dosage of 360 mg sustained-release (SR) verapamil given every 12 hours concomitantly with rifampin achieved median verapamil exposures of 903.1 ng.h/ml (AUC 0-12h), similar to those in persons receiving daily doses of 240 mg verapamil SR but not rifampin. Norverapamil:verapamil, R:S verapamil and R:S norverapamil AUC ratios were all significantly greater than those of historical controls receiving SR verapamil in the absence of rifampin, suggesting that rifampin administration favors the less-cardioactive verapamil metabolites and enantiomers. Finally, rifampin exposures were significantly greater after verapamil administration. Our findings suggest that a higher dosage of verapamil can be safely used as adjunctive treatment in rifampin-containing treatment regimens.

## INTRODUCTION

The need for lengthy TB treatment is attributed to antimicrobial tolerance of *Mycobacterium tuberculosis* (Mtb) (1), suggesting the potential of therapeutic approaches specifically targeting tolerant Mtb. Within days of infecting macrophages, Mtb subpopulations become tolerant to rifampin and other anti-tubercular drugs due to the activity of bacterial efflux pumps (2–4). Accordingly, Mtb strains with mutations in the Tap/Rv1258c efflux pump, which is induced upon macrophage infection (5), do not develop macrophage-induced rifampin tolerance (2, 4, 6). Beyond mediating rifampin tolerance, Tap/Rv1258c also promotes Mtb growth in macrophages in the absence of antibiotics (4). *Rv1258c* expression is induced in Mtb in sputum from patients being treated with a rifampin-containing regimen, supporting its relevance in TB (7).

Drugs with known activity against bacterial efflux pumps have also been shown to inhibit Mtb rifampin tolerance as well as growth within macrophages (3, 4, 6, 8). Among the most active of these is verapamil, which has been in clinical use worldwide for decades and is on the World Health Organization’s list of essential medicines (9). Its pharmacology and adverse effect profile are well-characterized, and it is available as a generic medication (10). Verapamil has been successfully used for both cardiovascular and non-cardiovascular indications (11, 12) and appears not to have a major effect on blood pressure in non-hypertensive populations (13). Animal studies show that verapamil is highly concentrated in tissue, including lung, with concentrations 40-fold or higher than those in plasma (14, 15). Along with inhibiting Mtb drug efflux, verapamil may potentiate Mtb killing through inhibitory effects on mammalian transporters (8, 16). In support of these findings, mice infected with drug-sensitive Mtb and given shorter courses of a rifampin-containing regimen and adjunctive verapamil had increased rates of relapse-free cure (17). Moreover, calcium channel blocker use in humans has been associated with a reduced incidence of TB (18).

While verapamil is useful in the management of cardiovascular disease due to its calcium channel antagonism, it also inhibits P-glycoprotein (Pgp). Verapamil is administered as a racemic mixture; both its major metabolite norverapamil and its R-enantiomer have substantially-reduced cardiac activity (19–21), but similar Pgp inhibitory activity. Prior work has shown that verapamil inhibits Mtb rifampin efflux through its Pgp inhibitory activity and that both R-verapamil and norverapamil have similar efficacy as racemic verapamil in inhibiting Mtb rifampin efflux, macrophage-induced drug tolerance and intramacrophage growth (4, 6). Notably, norverapamil is present at plasma levels similar to or higher than verapamil, potentially augmenting the effects of verapamil on rifampin efflux (22–27).

The major barrier to evaluation of verapamil as an adjunctive TB therapy is its greatly increased metabolism induced by rifampin, resulting in very low serum verapamil concentrations (28). We therefore sought to determine through a dose-finding pharmacokinetic study of verapamil given to patients with TB receiving rifampin-based therapy whether we could determine a compensatory dose increase of verapamil to offset its increased metabolism caused by rifampin. Our secondary goals were to determine the safety and tolerability of verapamil given in this way to patients with TB without known cardiac disease and to determine concentrations of verapamil enantiomers and norverapamil during rifampin-based TB therapy.

## METHODS

The most appropriate pharmacokinetic (PK) parameter to consider for an efflux pump inhibitor is uncertain. We chose an AUC (0-12h) target of 1000 ng.h/mL which is comparable to or lower than AUC ranges for human patients taking well-tolerated, moderate doses of sustained-release verapamil (240mg/day) in the absence of rifampin (29, 30).

Study participants with smear-positive pulmonary tuberculosis were enrolled from the National Institute for Research in Tuberculosis (NIRT) and Kilpauk Medical College and Hospital in Chennai, India along with the Regional Medical Research Center and SCB Cuttack Medical College and Hospital in Bhubaneswar, India (Figure 1). All participants were 18 to 55 years old and weighed 45-75 kg. They were in their last week of first-line TB therapy with daily rifampin, isoniazid (INH), and ethambutol per India’s National TB Elimination Programme (NTEP), had converted sputum smears to negative, and were clinically improved. TB drug dosing was based on NTEP standardized weight bands. All TB medications were provided from the NTEP. Verapamil was provided as Calaptin SR (Abbott) and obtained from commercial suppliers. We chose the extended-release formulation of verapamil since the less frequent dosing simplifies treatment and provides for more consistent drug levels (31).

**Figure 1:**
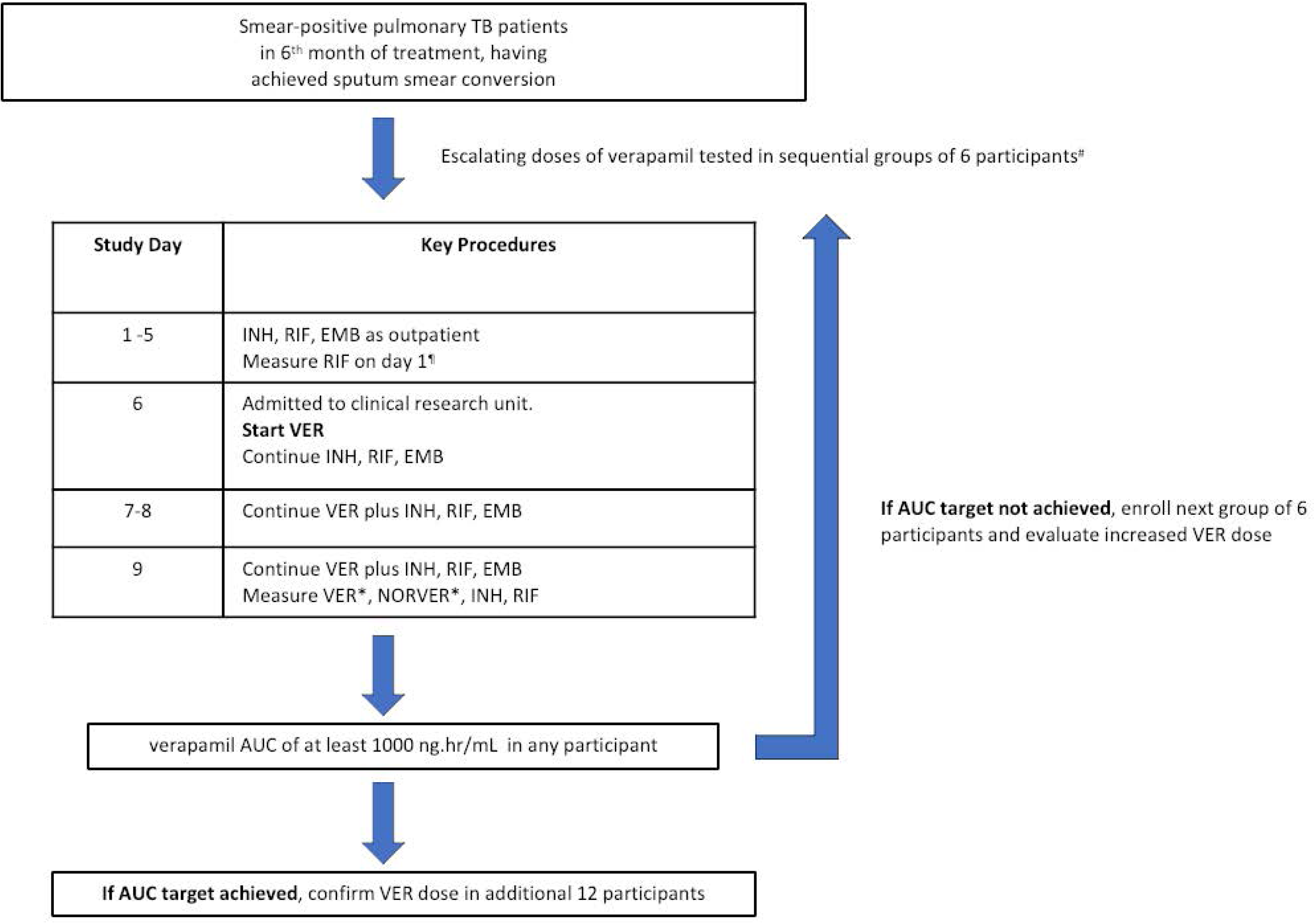
Study procedures

After providing written informed consent, participants underwent physical examination and baseline testing including complete blood count, renal and liver function testing, HIV serology, urine pregnancy testing, chest radiography and electrocardiography (EKG). Exclusion criteria are detailed in Supplementary Table S1.

Escalating dosages of sustained release verapamil were studied in sequential groups of 6 study participants. The first dose group included only 3 participants as we anticipated very low verapamil levels in this dose range. Blood was collected for measurement of rifampin levels on study day 1. Participants received rifampin, INH, and ethambutol under direct supervision at the study site outpatient clinic for study days 1-5. Participants were subsequently hospitalized to facilitate intensive PK monitoring. On study day 6, sustained-release verapamil was co-administered with rifampin, INH, and ethambutol prior to meals. All medications were given under direct supervison, and verapamil doses were administered 12 hours apart. Participants underwent daily EKGs from study days 6-9. Pulse and blood pressure were checked before each verapamil dose. Individuals with PR interval greater than 200 milliseconds, pulse less than 55 beats per minute or systolic blood pressure less than 90 mm Hg had verapamil held and the measurements repeated in 1 hour; if the abnormal value resolved then verapamil was given but if not, then the subject was excluded from further participation in the study.

On study day 9, blood was collected for measurement of INH, rifampin, verapamil and norverapamil levels prior to morning medication administration and then at 1, 2, 4, 8, and 12 hours afterward. Drug levels were measured for each group of 6 participants and reviewed by the Data Safety and Monitoring Committee before advancing to the next dose in another group of 6 participants. The study protocol dictated that verapamil dose escalation would be halted once an AUC (0-12h) of at least 1000 ng.h/mL was achieved in any participant, or if any participant experienced an adverse effect of grade 3 or higher as defined by the Division of AIDS, NIAID (32) or developed Mobitz type II or complete heart block. Once the AUC target had been achieved, the verapamil dose was studied in a confirmatory group of 12 participants.

### Laboratory methods

Drug levels were measured in parallel in two separate laboratories, NIRT and SITEC Labs. The NIRT laboratory measured racemic verapamil and norverapamil, as well as INH and rifampin using high pressure liquid chromatography. The SITEC laboratory measured R-verapamil, S-verapamil, R-norverapamil and S-norverapamil using liquid chromatography tandem mass spectrometry. The SITEC laboratory results are used in this report for verapamil and its metabolites because only that laboratory measured enantiomer concentrations. The NIRT laboratory results are used in this report for INH and rifampin levels. Sample collection and processing details, analytical methods, and SNP genotyping methods are described in the Supplementary Information.

### Data analysis

Study data were doubly entered into a Promasys database (Omnicomm, Ft. Lauderdale, Florida). Pharmacokinetic analyses were done with WinNonlin Version 8.1 (Certara, Princeton, NJ) using noncompartmental analyses. Data displays, descriptive statistics and AUC determinations were performed using Prism 9 (GraphPad, San Diego, Calif.); validation of the accuracy of the AUC (0-t) calculations using Prism was by comparison with WinNonlin outputs using Bland Altman testing, that showed a 5% difference (bias 5.5%, SD of bias 4.7%, 95% limits of agreement -3.5 to 14.7%). To allow comparison of the verapamil AUC (0-12h) with historical controls reporting AUC (0-24h), the calculated individual subject AUCs (0-12h) were doubled because the subjects were at steady state and receiving verapamil every 12h. The same doubling of the AUC (0-12h) to estimate the AUC (0-24h) was performed for data from Mattila et al (23), for identical reasons. Subjects were not included in the final analyses if there were missing data.

Because the published data for historical controls did not include raw data to allow the calculation of the variance of reported ratios, and attempts to obtain these data from study authors were unsuccessful, we used Bayesian methods to estimate the population distributions and ratios of both historical data and our data. The Bayesian analyses were conducted using Matlab (Mathworks, Natick, MA) as detailed in the Supplemental Information Statistical Appendix 1 and elsewhere (33).

### Ethics review and approval

This study underwent ethics review by the human subjects research committees at the National Institute for Research in Tuberculosis and Kilpauk Medical College and Hospital in Chennai, India along with the Regional Medical Research Center and SCB Cuttack Medical College and Hospital in Bhubaneswar, India. Each site provided ethical approval. The study was authorized by the Office of Drugs Controller General (India, FTS 35508). This trial was registered at the Clinical Trials Registry – India (CTRI/2016/05/006928) (34).

## RESULTS

No serious adverse events attributable to verapamil were observed in the study, including in the highest dosage group. In no case was a participant’s verapamil dosage held due to an abnormal EKG.

The detailed NIRT and SITEC data are shown in the Supplement (Figures S1 and S2); there was good agreement between the NIRT and SITEC results for total verapamil and norverapamil. Verapamil exposures in the first two verapamil dosage groups were low, with a median AUC(0-12h) of 60.5 ng.hr/mL (IQR 38.5 – 102) in the 120 mg twice daily group and a median AUC(0-12h) of 349 ng.hr/mL (IQR 319.5 – 445.5) in the 240 mg twice daily group (Supplementary Table S2). At a verapamil dosage of 360 mg twice daily, we attained the pre-specified AUC(0-12h) target (>1000 ng.h/mL) among 2 of 6 participants and subsequently advanced this dosing into a larger confirmatory group of 12 participants. The 18 participants receiving the 360 mg twice daily verapamil had a median age of 30 years (IQR 24 – 42), median BMI of 19.2 kg/m^2^ (IQR 17.7 – 21.9) and received a median of 9.65 mg/kg (IQR 9.3 – 10) of rifampin daily (Table 1). The median, arithmetic mean and geometric mean of the verapamil AUC (0-12h) for the 16 subjects with complete data were 903, 1020 and 835 ng.h/ml, respectively, with an IQR of 443 to 1298. Results for other analytes are shown in Table 2. When adjusted to AUC(0-24h) to allow for comparisons with previously published data, these values were comparable to steady state values reported for individuals taking 240mg verapamil SR once daily or 120 mg twice daily in the absence of rifampin (Figure 2A) (23, 25, 26, 29, 35).

**Figure 2.**
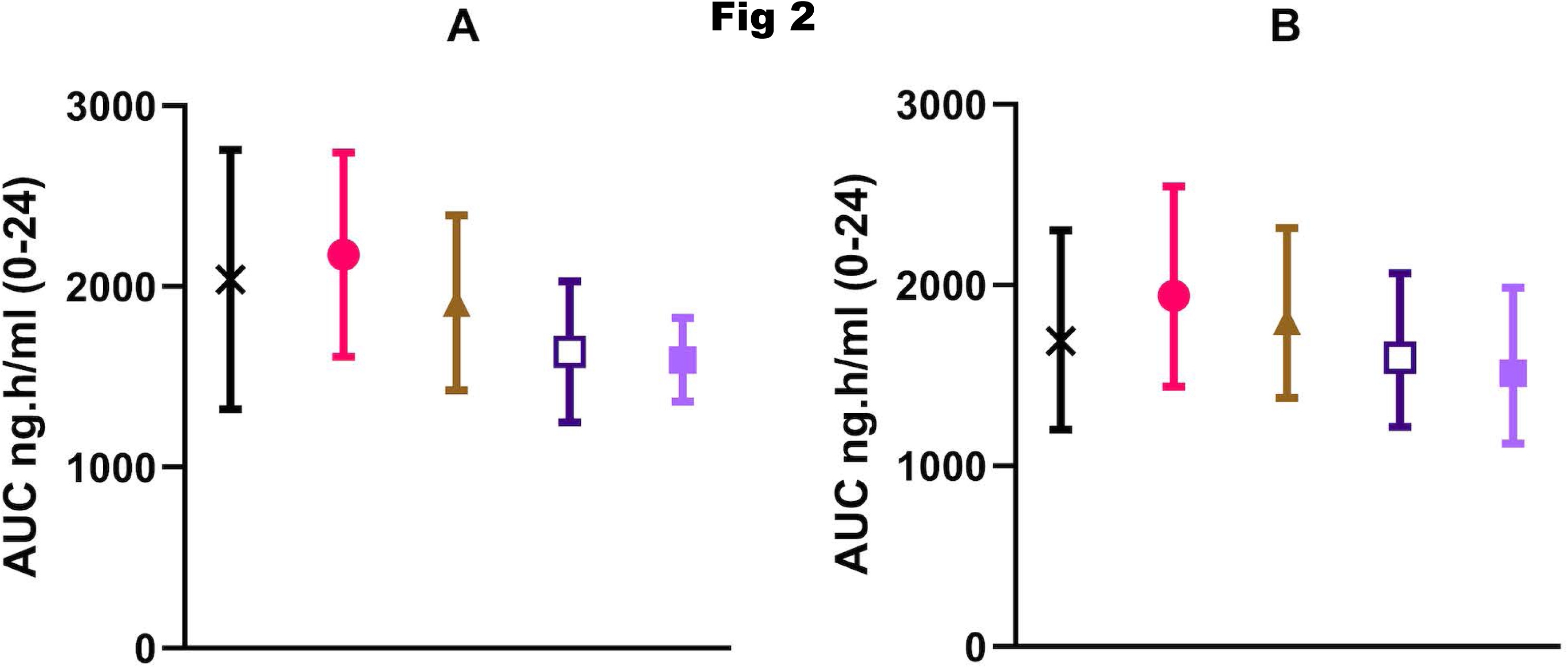
2A. Verapamil AUC (0-24h) for current study and historical comparators. Observed data are displayed as mean, and 95% frequentist confidence interval of the mean assuming a Gaussian distribution, with the current study and Mattila et al data (23) adjusted from the respectively measured or reported AUC (0-12h) to estimate AUC (0-24h). Reading from left to right: current study (n=16), black with X; Mattila et al. (n=12), filled red circle; Hla et al. (29) (n=10), brown triangle; Abernethy et al. (25) (n=8), dark purple hollow square, Lemma et al. (26) (n=12), light purple filled square. This is provided for easy comparison with the literature, acknowledging that the reported 95%CI values are potentially incorrect because of the assumption of a Gaussian data distribution. Details of comparison studies are provided in Supplementary Table S3. **2B. Verapamil AUC (0-24h) Bayesian population estimates** The posterior arithmetic mean of the geometric mean over the population is shown, along with the 95% Bayesian confidence interval for the geometric mean over the population, under the assumption (justified in Statistical Appendix 1 section 4.1) that the population distribution is log-Gaussian. The current study value is not significantly different from that of each of the other studies (< 0.70) (see Statistical Appendix 2 item 6 and Table S4a). Details of comparison studies are provided in Supplementary Table S3.

**Table 1:** Demographic characteristics of study participants receiving verapamil SR 360mg twice daily

| Gender | Age Range (yr) | Weight (kg) | Height (m) | BMI | Study Phase |
| --- | --- | --- | --- | --- | --- |
| Female | 41-45 | 47.5 | 1.68 | 16.8 | Initial group (n=6) |
| Male | 51-55 | 45 | 1.70 | 15.6 | Initial group |
| Male | 20-25 | 61.8 | 1.75 | 20.2 | Initial group |
| Male | 26-30 | 49.6 | 1.62 | 18.9 | Initial group |
| Male | 36-40 | 52.7 | 1.68 | 18.7 | Initial group |
| Male | 31-35 | 46.2 | 1.66 | 16.8 | Initial group |
| Male | 26-30 | 61.2 | 1.70 | 21.2 | Confirmatory group (n=12) |
| Male | 20-25 | 48.4 | 1.78 | 15.3 | Confirmatory group |
| Male | 36-40 | 53.5 | 1.74 | 17.7 | Confirmatory group |
| Male | 31-35 | 62.6 | 1.67 | 22.4 | Confirmatory group |
| Female | 26-30 | 46 | 1.45 | 21.9 | Confirmatory group |
| Male | 20-25 | 54.2 | 1.57 | 22.0 | Confirmatory group |
| Male | 41-45 | 64.6 | 1.65 | 23.7 | Confirmatory group |
| Male | 41-45 | 48 | 1.58 | 19.2 | Confirmatory group |
| Male | 20-25 | 45.1 | 1.48 | 20.6 | Confirmatory group |
| Male | 46-50 | 48 | 1.48 | 21.9 | Confirmatory group |
| Female | 20-25 | 55 | 1.72 | 18.6 | Confirmatory group |
| Male | 41-45 | 51 | 1.42 | 25.3 | Confirmatory group |

Bayesian analysis confirmed that the current study verapamil AUCs were not significantly different from those of each of these historical controls (Figure 2B and Table S4a). For statistical definitions and precise meanings of abbreviated phraseology for these and all other Bayesian analyses, see Statistical Appendix 2.

We measured levels of verapamil enantiomers and norverapamil and determined their relative concentrations by calculating the ratios of the AUCs of norverapamil to verapamil, R-verapamil to S-verapamil and R-norverapamil to S-norverapamil (Table 2 and Figure 3). These data are reported as described in Statistical Appendix 2 (item 11). The norverapamil:verapamil AUC ratio of our study participants (2.11, 1.89-2.35) was significantly greater (>0.97) than the ratio in each study conducted among participants receiving 240 mg SR verapamil at steady state not taking rifampin (23–26) (Figure 3a, Table S4b). We note that Barbarash et al. (28) observed an even larger norverapamil: verapamil ratio when verapamil was co-administered with rifampin, perhaps because their measurements were made after a single dose of 120 mg immediate release verapamil when steady state levels would not have been achieved. Together these findings suggest that rifampin co-administration increases the relative concentrations of norverapamil to verapamil, even before steady-state levels are achieved and maintains these higher relative levels under steady state concentrations.

**Figure 3.**
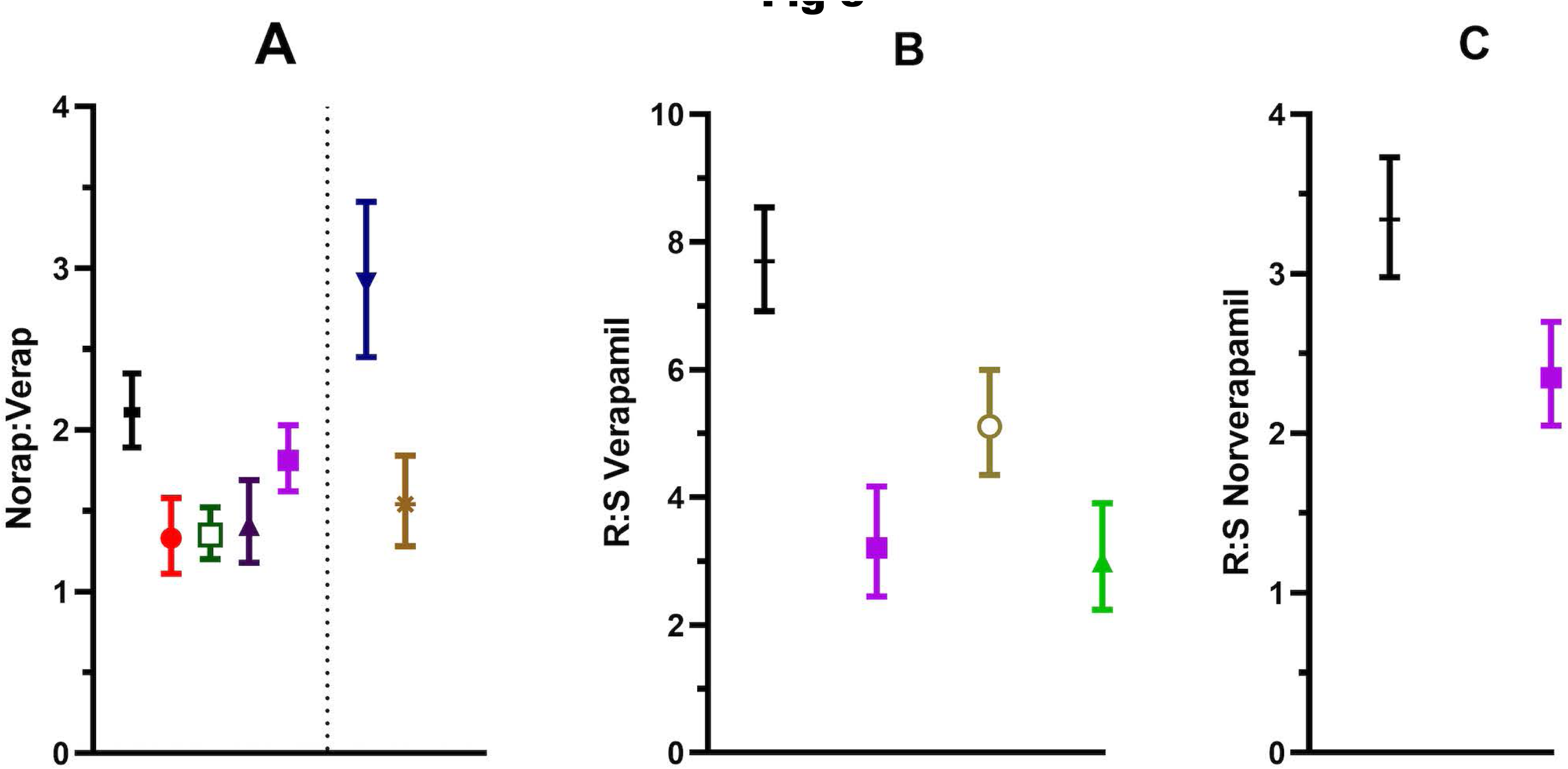
3A. Norverapamil:verapamil AUC ratios for current study and historical comparators. Reading from left to right, current study, black horizontal line (N= 16); Mattila et al. (23) (n=12), filled red circle; Norris et al. (24) (n=22), green hollow square; Abernethy et al. (25) (n=8), dark purple triangle; Lemma et al. (26) (n=12), light purple solid square. Separated from these by a vertical dotted line are Barbarash et al. (28) post rifampin (n=6), dark blue downward triangle; and Barbarash et al. pre-rifampin, brown asterisk. The Barbarash data are separated because this was a single immediate release verapamil dose study, in contrast to the other studies which were all steady state delayed release verapamil studies. Results are reported as described in the Statistical Appendix 2 (item 11). The probabilities that the present study value is greater than that of each other study are > 0.999 except for Lemma et al (0.973), Barbarash post-rifampin (0.003) and pre-rifampin (0.996). The probability that Barbarash et al. post-rifampin is greater than pre-rifampin is 0.9999. Details of comparison studies are provided in Supplementary Table S3. **3B. R:S verapamil AUC ratios for current study and historical comparators** Reading from left to right, current study, black horizontal line (N=16); Lemma et al. (26) (n=12), purple solid square; Fromm et al. (36) (n=8) pre-rifampin, beige hollow circle; Fromm et al. post-rifampin, green triangle. Results are reported as described in the Statistical Appendix 2 (item 11). The present study value is significantly greater than that of each other study’s value (>0.999). The probability that Fromm et al. pre-rifampin is greater than post-rifampin is 0.999. Details of comparison studies are provided in Supplementary Table S3. **3C. R:S norverapamil AUC ratios.** Reading from left to right, current study, black horizontal line (N=18) and Lemma et al. (26), purple solid square. Results are reported as described in the Statistical Appendix 2 (item 11). The Bayesian probability that the current study value is greater than that of Lemma et al is 0.999. Details of comparison studies are provided in Supplementary Table S3.

**Table 2:**
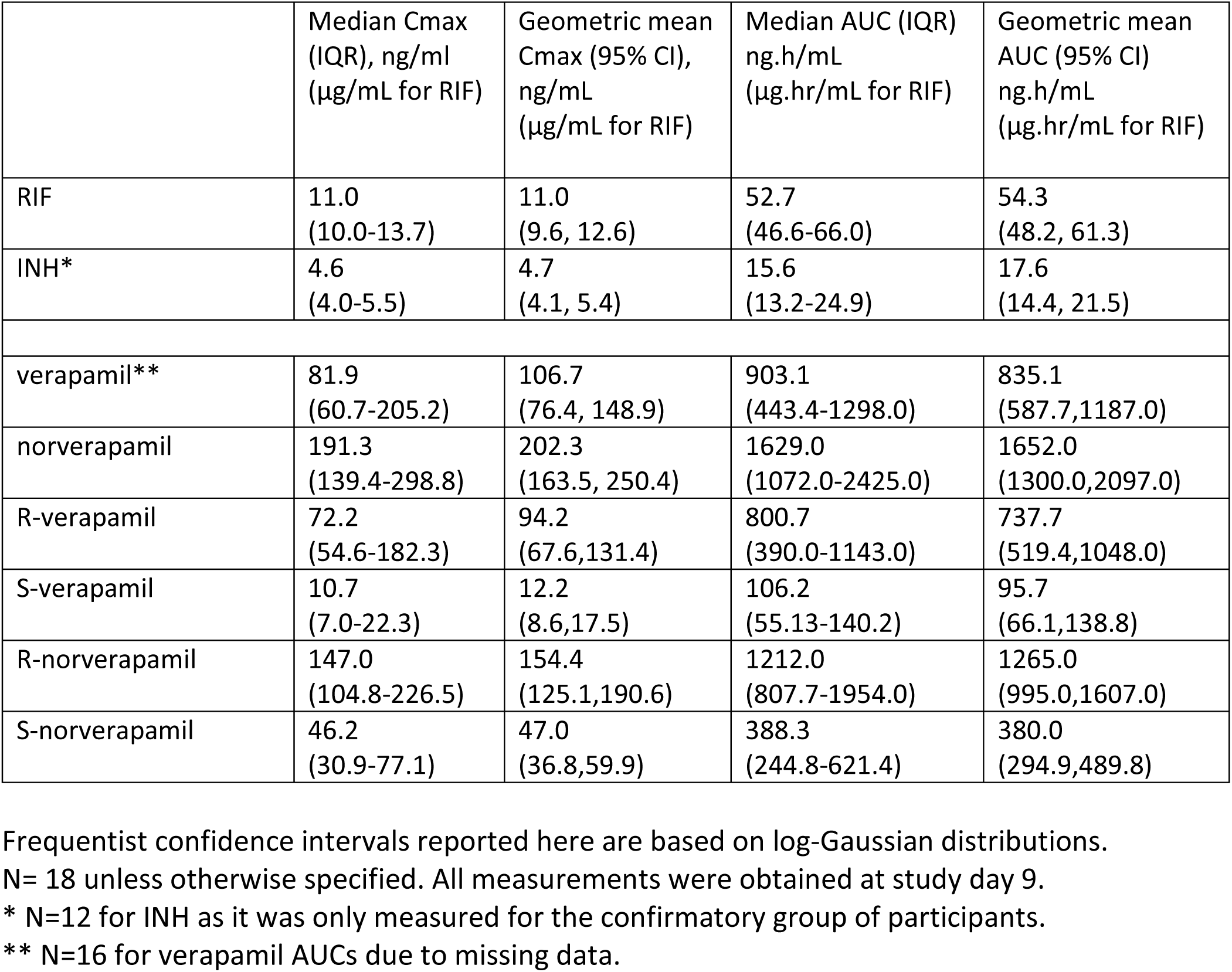
Summary PK data among participants receiving verapamil SR 360 mg twice daily.

The ratio of the AUC of R-verapamil to S-verapamil (7.70, 6.92-8.54) was also significantly higher than that reported for persons receiving verapamil in the absence of rifampin (Figure 3b, Table S4c) (26, 36). These results were different from those reported by Fromm et al., where the R:S ratio was significantly (0.999) lower among persons taking verapamil with rifampin than in those not taking rifampin (36). This difference could reflect differences in the study population. We note that the results in our study were internally consistent: R-norverapamil to S-norverapamil ratios (3.34, 2.98-3.73) were also significantly greater than those reported in participants receiving verapamil in the absence of rifampin (Figure 3c, Table S4d) (26). As previously reported (27), the R:S ratios for both verapamil and norverapamil decreased over time following administration of the last verapamil dose, while remaining greater than 1 (Figure 4).

**Figure 4.**
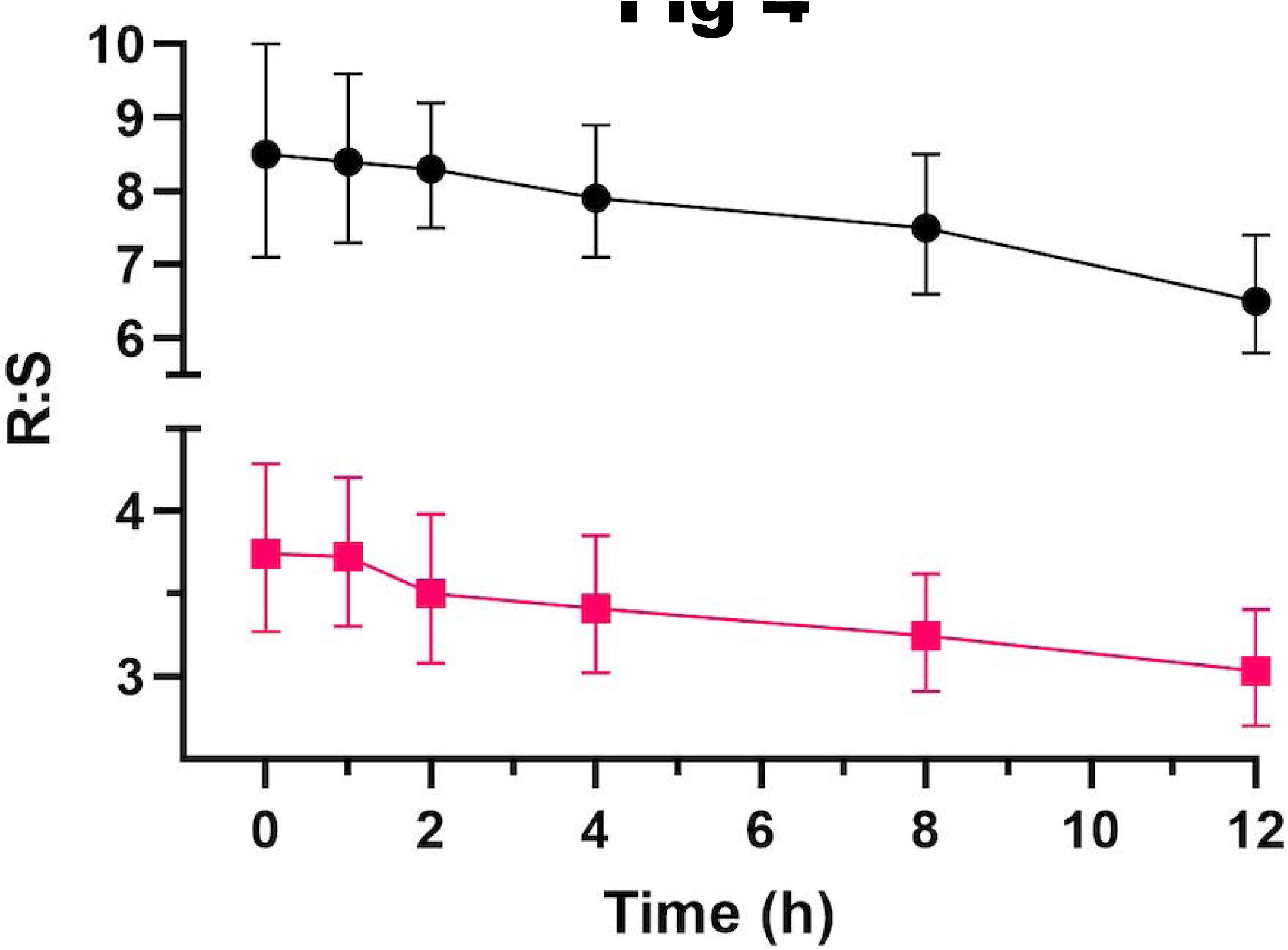
Time trends in R:S enantiomer ratios of verapamil and norverapamil. Verapamil and norverapamil R:S ratios over the time of administration of the 7^th^ oral dose of verapamil SR 360mg given every 12h to subjects who were also receiving rifampin. Sample geometric means and 95% CI of the geometric means of the ratios of the plasma levels of the drugs are shown. Red squares, R:S norverapamil, N=18; black circles R:S verapamil, N=16; the lower N for verapamil is because of undetectable S enantiomers or missing values.

We asked if verapamil AUCs were altered by genetic variants in P-glycoprotein (encoded by *ABCB1*) which are associated with alterations in verapamil transport (37, 38). Individuals homozygous for both minor alleles of two closely linked single nucleotide polymorphisms (SNPs) in *ABCB1* are reported to have increased serum verapamil levels (38). One of these, rs1045642, is a synonymous change (NM_001348944.2:c.3435T>C) in exon 26, and the other, rs2032582, is a nonsynonymous change (NM_001348946.2:c.2677T>G), in exon 21 (Ser893Ala). We examined the effect of the two SNPs individually on verapamil AUC, as described (33). rs1045642 was associated with differences; both CC individuals had higher verapamil AUCs than did the TT and TC subjects (Figure 5A, and Supplementary Table S4e). For rs2032582, only one of the two GG homozygotes had elevated verapamil AUCs; this individual was also CC homozygous at rs1045642 (Figure 5B). Together, these findings suggest that ABCB1 rs1045642 is the primary driver of the differences in verapamil metabolism observed in the combined haplotype (38). A genetic variant in the cytochrome P450 enzyme encoded by *CYP3A5*, which metabolizes verapamil, is also reported to influence the verapamil AUC (39). *CYP3A5* rs776746 (NM000777.5(CYP3A5):c.219-237A>G) in intron 3 encodes the nonfunctional CYP3A5*3 allele (39). Minor allele GG homozygotes expressing the truncated form are reported to be non-expressors and are associated with higher verapamil levels (37). However, we did not find significant differences in verapamil AUCs based on rs776746 genotype in this study (Figure 5C). This may be due to small sample size, differences in study populations, and/or the inductive effects of rifampin on verapamil that outweigh any differences that would otherwise be seen due to variable CYP3A5 activity.

**Figure 5.**
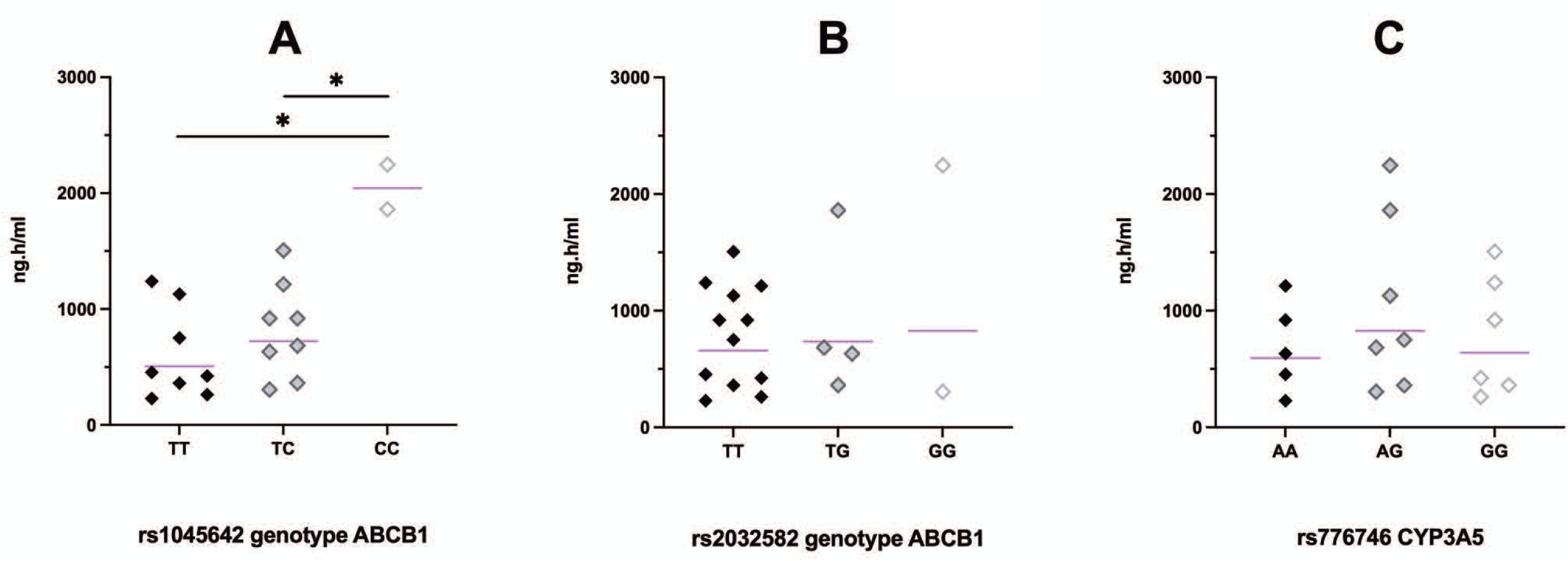
Comparison of verapamil AUC grouped by SNP genotype. Verapamil AUC (0-12h), ng.h/ml, for each study participant, grouped by genotype at (A) rs1045642, ABCB1 3435T>C; (B) rs2032582, ABCB1 2677T>G; and (C) rs776746, CYP3A5 6986A>G. Asterisks indicate that the restricted geometric mean of the CC allele in figure A is significantly greater than each of the two alternative alleles (see Statistical Appendix 2, item 13). Unspecified comparisons have probabilities of ≤0.91. Bars indicate geometric means.

Because verapamil is reported to increase rifampin exposure in mice (40, 41), we examined the effect of verapamil on rifampin AUCs in those ten subjects who had both pre-and post-verapamil data. We found that there was a significant increase (1.5 fold) in rifampin AUCs after three days of verapamil treatment (Figure 6). In contrast, INH AUCs were not significantly increased by verapamil.

**Figure 6.**
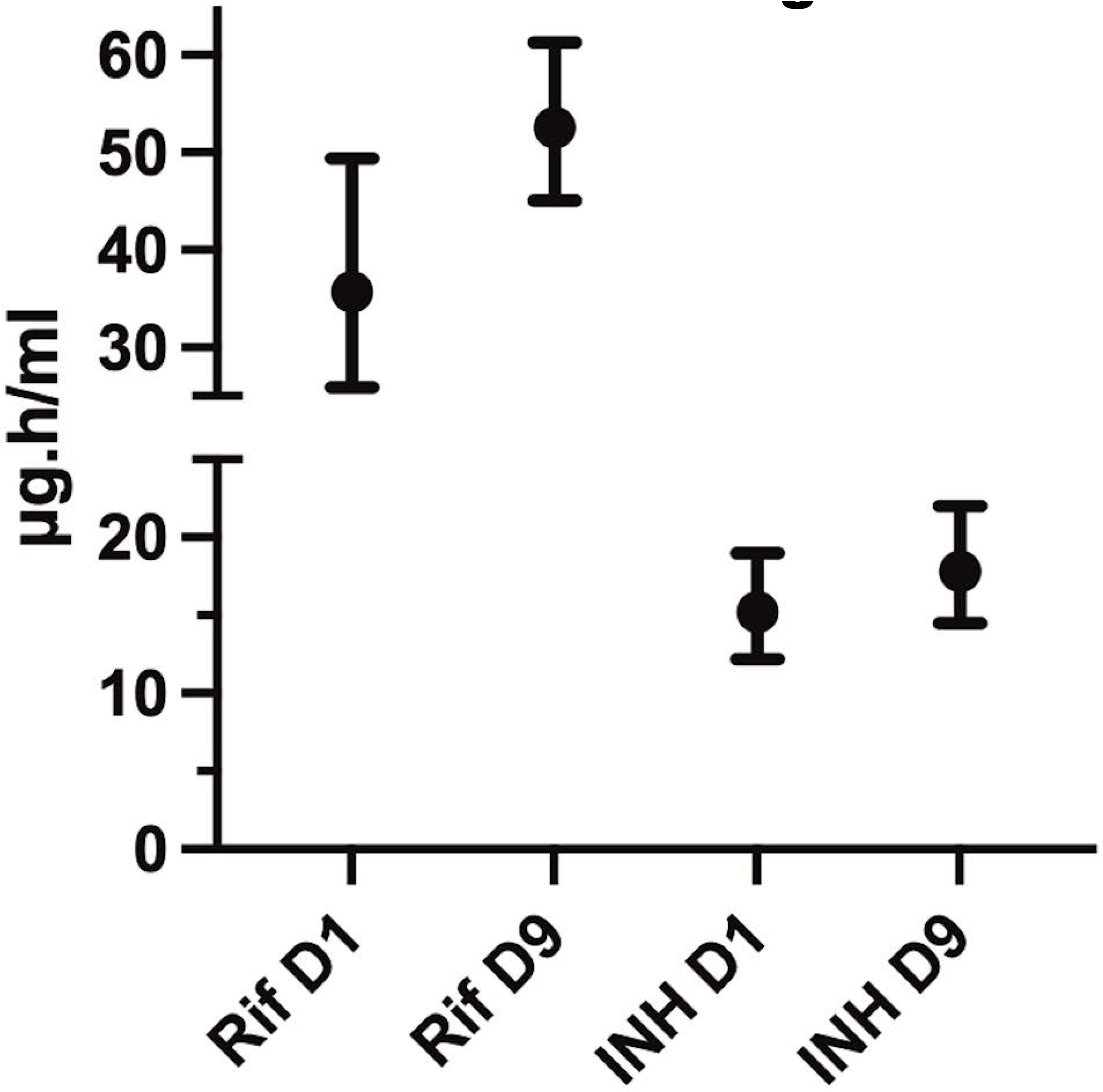
Effect of verapamil on RIF and INH levels. Comparison of INH and rifampin AUCs (0-8h) in paired subjects (N=10) before and after receiving 7 doses (84 h) of twice daily verapamil (360 mg SR q 12h), showing the effect of verapamil administration on INH and rifampin exposures. Rif D1, rifampin AUC prior to receiving verapamil; Rif D9, rifampin AUC after receiving 7 doses of verapamil; INH D1, INH AUC prior to receiving verapamil; INH D9, INH AUC after receiving 7 doses of verapamil. Results are reported as described in Statistical Appendix 2, item 11. The Bayesian probabilities of the geometric mean drug exposures while receiving verapamil being greater than the geometric mean drug exposures prior to receiving verapamil in these paired subject analyses are 0.98 for rifampin and 0.90 for INH.

## DISCUSSION

In this investigation, we studied escalating doses of verapamil given to participants receiving rifampin-based TB therapy to determine whether increased verapamil clearance can be offset by higher verapamil doses. We found that treatment with verapamil 360mg SR twice daily can attain verapamil exposures similar to those among persons taking moderate doses of verapamil (240mg SR daily) in the absence of rifampin (29, 30).

As in previous studies, we found that R-verapamil is the predominant verapamil enantiomer in blood. Moreover, norverapamil to verapamil ratios were significantly increased in the presence of rifampin. This is likely due to the selectively increased metabolism of verapamil versus norverapamil (28). Thus, verapamil metabolites and enantiomers may contribute significantly to the effects of verapamil in inhibiting efflux and may potentially allow for further increasing verapamil doses, without increasing cardiotoxic potential, in rifampin-containing regimens.

We also considered the potential of verapamil to increase rifampin levels as increased rifampin levels are associated with therapeutic benefit in TB (42). It is difficult to predict the effects of verapamil on rifampin levels, since verapamil not only inhibits CYP3A4, but also inhibits gut P-glycoprotein (Pgp) with short term administration and induces Pgp with longer term administration (26, 43). The rifampin dosing in this study was consistent with current practice and achieved rifampin exposures similar to those reported in other studies where verapamil was not administered (44). We observed a statistically significant, but probably not a clinically significant, increase in rifampin exposures after verapamil treatment, suggesting that verapamil enhances rifampin absorption similar to what is seen with rifampin-piperine coadministration (45). It is possible that longer term verapamil administration could alter these results if gut Pgp was not fully induced by less than a week of verapamil administration.

Verapamil remains of interest as an adjunctive therapy even with new TB regimens coming into use. Although rifapentine and fluoroquinolone-based therapy permits treatment shortening to 4 months in drug-susceptible TB (46), adjunctive efflux inhibitory treatment might permit further shortening given macrophage-induced fluroquinolone and rifamycin tolerance (3). Verapamil inhibits macrophage-induced rifabutin tolerance and is likely to inhibit rifapentine tolerance as well ((3) and Adams KN, unpublished data 2023).

Notably, no major adverse events were noted even in those subjects with the highest measured verapamil exposures. At the same time, rifampin and verapamil combination regimens carry the risk of substantial increases in verapamil levels if rifampin alone is discontinued. This risk is mitigated by two findings: 1) verapamil doses of 960 mg daily or greater have been safely used in the absence of rifampin, suggesting a wide therapeutic window with verapamil (11, 12); 2) The delayed timing of CYP3A4 recovery provides a buffer against the effects of missed rifampin doses (36). Prolonged discontinuation of rifampin, but not verapamil, would result in substantially higher verapamil exposures and potential toxicity.

Use of a combined rifampin/verapamil formulation would mitigate this, as well as careful monitoring and counseling to ensure medication adherence. If verapamil is added to a rifamycin-containing treatment regimen then it should only be added after full CYP3A4 induction has occurred, around a week after starting the rifamycin (36).

This study has limitations that affect its generalizability. Study participants did not have major medical comorbidities or concomitant medication use. Women were underrepresented, despite enrollment efforts. The number of subjects studied was relatively small, especially for the before-after rifampin and INH exposure studies and genetic association studies, requiring confirmatory studies in other populations. The use of historical controls of verapamil, and verapamil metabolite exposures, rather than comprehensive before-after studies in the same population requires confirmation of these results in future studies with same-population controls. Our study population was carefully selected and intensively monitored during drug administration, limiting generalizability about safety in a less well selected and monitored population. While intensive cardiac screening with echocardiology and electrocardiograms would be difficult to implement in less-resourced settings, careful clinical examination might be adequate to identify persons who should not receive adjunctive verapamil. We note that verapamil has been widely used for noncardiac indications without intensive cardiac monitoring and has been well tolerated.

In summary, verapamil is appealing to study for adjunctive TB treatment given the multiple potential pathways through which it may increase anti-tubercular drug efficacy and counter TB pathogenesis, and its well characterized pharmacology and decades of clinical experience. We have established a well-tolerated, compensatory dose of verapamil to help inform future studies of adjunctive verapamil in the context of rifampin-based TB therapy.

## STUDY HIGHLIGHTS

### • What is the current knowledge on the topic?

Verapamil metabolism is greatly accelerated during rifampin administration, limiting the potential use of adjunctive verapamil therapy in patients with TB being treated with rifampin. Such treatment has the potential to shorten TB therapy, and to reduce the potential for emergence of drug resistance, both related to the ability of verapamil to decrease TB drug tolerance.

### • What question did this study address?

This study sought to determine a compensatory dosing strategy of verapamil to offset its increased metabolism when given to patients with TB receiving rifampin-based therapy.

### • What does this study add to our knowledge?

Verapamil sustained release 360mg twice daily achieved a similar drug exposure as compared to prior studies of verapamil 240mg once daily given to persons without TB. Verapamil co-administration appeared to increase the relative concentrations of the less cardioactive metabolites and enantiomers.

### • How might this change clinical pharmacology or translational science?

The results of this study are a proof of concept that compensatory dosing of verapamil can be achieved in the context of rifampin-based TB therapy and is well-tolerated.

## Data Availability

All data produced in the present work are contained in the manuscript

## ACKNOWLEDGEMENTS

We thank the study participants and the clinical staff of the study sites. We thank Dr. Danny Shen for expert guidance relating to verapamil assay design, Dr. Yusuf Hamied and CIPLA for facilitating the collaborative work with SITEC laboratories, along with Dr.Rohit Sarin, ex-Director, NITRD, New Delhi; Dr.Govindharajalu, Professor of Medicine, Govt. Kilpauk Medical College and Hospital, Dr.Dasarathi Das, Scientist F, RMRC Bhubaneswar and Dr.Sushrita Mohanty, Consultant Medical Officer, SCB Cuttack, Odisha for assistance with the study.

## AUTHOR CONTRIBUTIONS

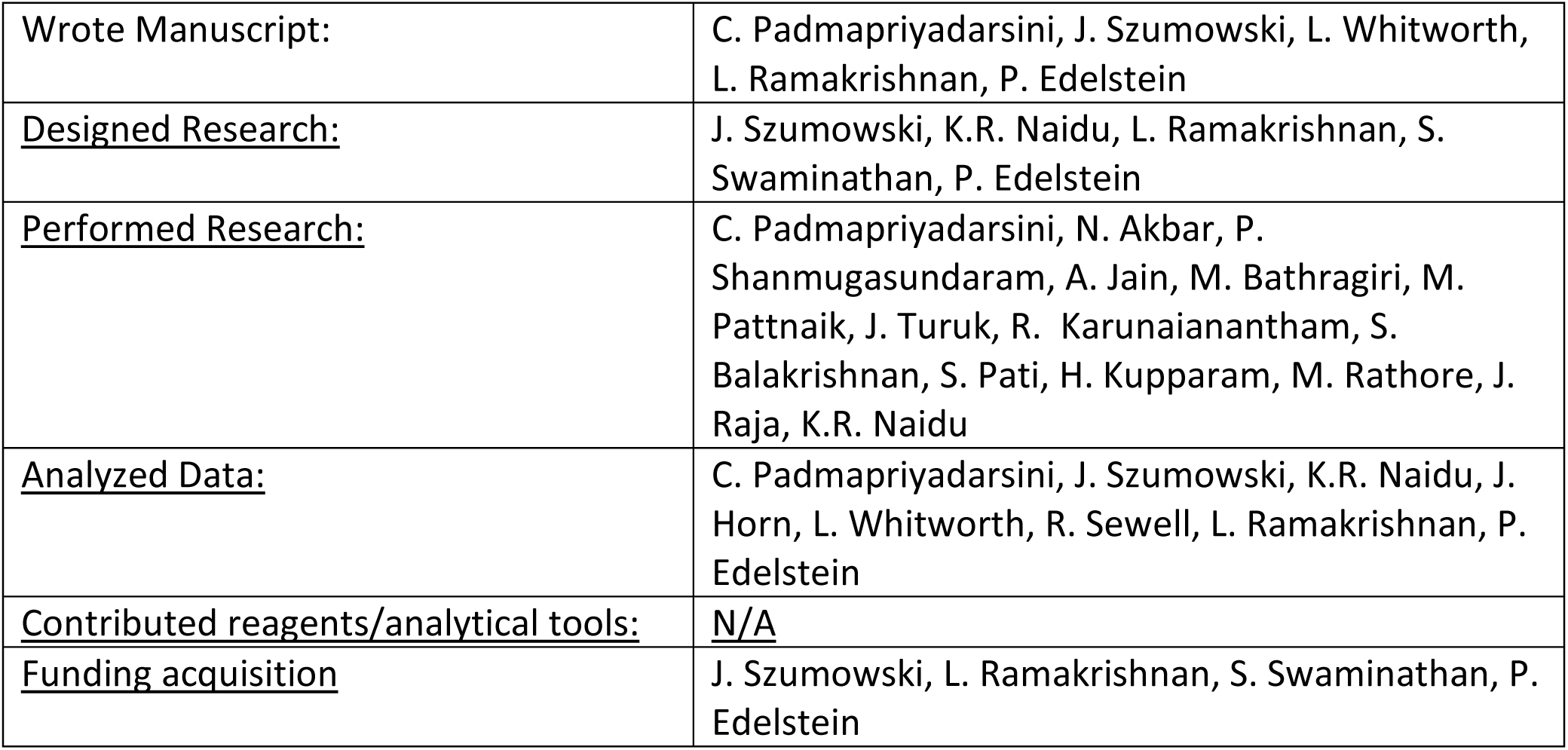

## Supplementary Information

### LABORATORY METHODS

Blood was drawn into Vacutainer lithium heparin 4 ml tubes (Becton Dickinson and Co., Franklin Lakes, NJ) and plasma was separated via centrifugation at 958 RCF for 5 minutes at room temperature. All specimens underwent centrifugation within 1 hour of phlebotomy. For each 1 ml of plasma, 10 microliters of 5% ascorbic acid solution were added for the rifampin assay plasma. All separated plasma was stored in two aliquoted cryogenic vials at -80°C, with one set of vials kept for analysis by SITEC Labs and the other for analysis by the NIRT laboratory. For genotyping assays, blood was collected into Vacutainer dipotassium ethylene diamine tetraacetic acid (K2EDTA) 4ml tubes (Becton Dickinson and Co., Franklin Lakes, NJ) and was stored in two aliquoted cryogenic vials at -80°C and shipped to NIRT. Samples were transported on dry ice. All samples were labeled only with coded identifiers.

#### NIRT methods

Drug levels were measured at NIRT via HPLC in batches. The analytical methods used for rifampin and isoniazid were described previously (1, 2). All reagents used were HPLC grade. Verapamil and norverapamil were obtained from Sigma Aldrich (≥99% purity). The plasma samples were deproteinized using 5% ammonia in water, vortex mixed for a minute and then centrifuged at 6720 RCF for 10 minutes at room temperature. The entire supernatant (approximately 550 μL) was loaded onto solid phase extraction cartridges (Oasis MCX 1cc 30mg, Waters Corporation, Milford, MA). The clear eluted filtrate (1 ml) was evaporated to dryness under nitrogen for approximately 30 minutes. The dried residue was dissolved in the mobile phase (75 µl), of which 20 µl was injected into the HPLC column (Ascentis Express C18, 15 cm X 4.6 mm ID, 2.7 µm particle size, Merck, Germany). An isocratic mobile phase was prepared by mixing 20 mM potassium dihydrogen orthophosphate adjusted to pH 2.5 with 85% orthophosphoric acid and acetonitrile in the ratio of 60:40 (v/v). Each chromatogram was run for 7 minutes at a flow rate of 0.8 ml/min at 40°C, with the PDA detector set at a wavelength of 200 nm. Data collection and acquisition were carried out using LabSolutions software version 5.92 (Shimadzu, Kyoto, Japan). The retention times for norverapamil and verapamil were 3.8 and 4.0 minutes respectively. Analyte concentrations were calculated from linear regression analysis of verapamil and norverapamil concentration curves that were created using pooled human plasma; linearity was verified using correlation coefficient estimates. The calibration curve was linear over a range of 50 to 1000 ng/ml for both verapamil and norverapamil. The accuracy of plasma verapamil and norverapamil concentrations ranged from 94% to 102 %. The extraction efficiency was 94 to 104%. The intra-and inter-day relative standard deviations were below 10%.

#### SITEC methods

Drug levels were measured at SITEC via liquid chromatography tandem mass spectrometry (LC-MS/MS) in one batch. A bioanalytical method was developed for simultaneous determination of R and S enantiomers of verapamil and norverapamil and will be described in detail in a subsequent manuscript. The analytical method is based on chromatographic separation of enantiomers of verapamil and norverapamil in a single run and their detection using triple quadrupole MS/MS technique (API 6500) in MRM mode. MRM transition 455.1 – 165.1 for verapamil (R & S), 441.2 – 165.2 for R-norverapamil and 441.2 – 165.1 for S-norverapamil were used. The solvents and reagents used were all HPLC grade. Acetonitrile and methanol were purchased from J.T.Baker (Darmstadt, Germany). Ammonium trifluoroacetate (ATFA) was purchased from Sigma Aldrich, n-hexane was purchased from Merck, diethyl ether was purchased from Rankem and purified water was generated by an in-house system (Thermo Scientific TKA). Standards were procured from Daicel Chiral Technologies Private Ltd (India).

The developed LC–MS/MS method provided good resolution between enantiomers. The chromatographic separation between enantiomers is achieved using chiral column Chiralpak® AGP 150 x 4.0 mm, 5µm with mobile phase comprising of 20mM ATFA (pH:6.2): acetonitrile: methanol (92:5:3). Extraction of analytes from plasma was performed using liquid-liquid extraction. A mixture of diethyl ether and n-hexane in 1:1 proportion was used as extraction solvent. Isotope labelled analogue, R-verapamil D6 HCl, S-verapamil D6 HCl, R-norverapamil D6 HCl and S-norverapamil D6 HCl were used as internal standards. The extraction efficiency (mean recovery) was 100.83%, 107.49%, 99.77% and 98.82% for R-verapamil, S-verapamil, R-norverapamil and S-norverapamil respectively. The resolution between R & S enantiomers of verapamil is 2.19 and between R & S enantiomers of norverapamil is 1.62. The area response was linear over concentration ranges of 10.0–1500 ng/mL for R-verapamil, 2.0–400 ng/mL for S-verapamil, 6.0–1000 ng/mL for R-norverapamil, 2.0–400 ng/mL for S-norverapamil. The method validation data are summarized in Table S5.

#### Genotyping

Genomic DNA was extracted from whole blood using a QIAcube automated nucleic acid extractor (QIAGEN, Germany) using a QIAamp blood DNA mini kit (QIAGEN, Germany). DNA purity and quantity were measured using a Nanodrop 2000 instrument (Thermo Fisher, USA). Each DNA sample was diluted to 20 ng/µl concentration before storage at -20°C. Genotypes of single nucleotide polymorphisms in *ABCB1*, rs1045642 (chr7:87509329) and rs2032582 (chr7:87531302), and in *CYP3A5*, rs776746 (chr7:99672916) (genomic coordinates from build GRCh38.p14) were determined using pre-validated Applied Biosystems TaqMan human SNP Genotyping Assays in a 7500 fast Real Time PCR machine (Applied Biosystems, USA). Alleles were called using Applied Biosystems SDS software. For quality control, DNA was re-extracted from 3 samples separately and included in each allelic discrimination assay.

**Table S1.** Study exclusion criteria

|  |
| --- |
| Systolic blood pressure less than 100 mm Hg or heart rate less than 60 beats per minute |
| Clinical signs or symptoms of congestive heart failure or cirrhosis |
| Treatment with medications other than isoniazid, rifampin, ethambutol and pyridoxine |
| Co-morbid conditions: HIV, cardiac arrhythmia or congestive heart failure, diabetes mellitus or other uncontrolled medical condition |
| Current illicit drug use or heavy alcohol use |
| Pregnancy or lactation |
| Conduction delay on electrocardiogram (PR interval greater than 200 milliseconds or QRS greater than 120 milliseconds) |
| Ejection fraction less than 45% |
| Laboratory criteria:<br>• hemoglobin less than 9 mg/dL<br>• creatinine clearance less than 50 ml/min<br>• aspartate aminotransferase or alanine aminotransferase greater than 2.5 times the upper limits of normal |

**Table S2.** Verapamil and Rifampin Levels Among Participants Receiving Verapamil SR 240 or 480 mg per day

| VER SR Dose | Median VER Cmax (IQR), ng/mL | Median VER AUC (IQR), ng.hr/mL | Median RIF Cmax (IQR), µg/mL | Median RIF AUC (IQR), µg.hr/mL |
| --- | --- | --- | --- | --- |
| 240 mg/day<br>(120mg q12 hours)<br>N=3 | 7 (5-17) | 60.5 (38.5-102) | 10 (9.04-10.93) | 40.26 (36.51-52.25) |
| 480 mg/day<br>(240 mg q12 hours)<br>N=6 | 50.5 (47-71) | 349 (319.5-445.5) | 9.32 (7.91-10.6) | 48.72 (43.88-55.3) |

VER: verapamil, RIF: rifampin

**Table S3.** Verapamil dosing in comparison studies cited in Figures 2 and 3

| Study | Verapamil dose | Duration of treatment prior to AUC measurement | Participants (n) | Study comments |
| --- | --- | --- | --- | --- |
| **<u>Abernethy (3)</u>** | 240 mg daily | 14 days | 8 | Only “young participant” group included |
| **<u>Barbarash (4)</u>** | 120mg IR as single dose | See comments | <u>6</u> | <u>Single dose verapamil given prior to and then following 15 days of rifampin</u> |
| **<u>Fromm (5)</u>** | 120mg twice daily | See comments | <u>8</u> | <u>Verapamil given for 4 days, then continued with rifampin for 12 days</u> |
| **<u>Hla (6)</u>** | 240mg daily | 10 days | 10 | -- |
| **<u>Lemma (7)</u>** | 240mg daily | 37 days | 12 | -- |
| **<u>Mattila (8)</u>** | 120mg daily | 5 days | 12 | -- |
| **<u>Norris (9)</u>** | 240mg daily | 7 days | 22 | -- |

Unless otherwise stated, all participants received sustained-release verapamil and had AUC measured at steady state

**Table S4:** Bayesian Analysis tables Please refer to table S3 for study details. **Table S4a: Probability table - Verapamil AUC versus historical controls (Supplement to Figure 2B)** Probability greater than 0.95 indicates that the verapamil geometric mean AUC in study 1 is significantly higher than that in study 2. The inverse situation, with probability <0.05, indicates that the verapamil geometric mean AUC in study 1 is significantly lower than that in study 2.

|  | **Study 2** |  |  |  |  |
| --- | --- | --- | --- | --- | --- |
| **Study 1** | *Current study* | Hla | Abernethy | Mattila | Lemma |
| *Current study* | 0.500 | 0.372 | 0.600 | 0.256 | 0.695 |
| Hla (6) | 0.629 | 0.500 | 0.747 | 0.349 | 0.824 |
| Abernethy (3) | 0.400 | 0.253 | 0.500 | 0.156 | 0.618 |
| Mattila (8) | 0.744 | 0.651 | 0.844 | 0.500 | 0.893 |
| Lemma (7) | 0.306 | 0.176 | 0.382 | 0.107 | 0.500 |

**Table S4b:**
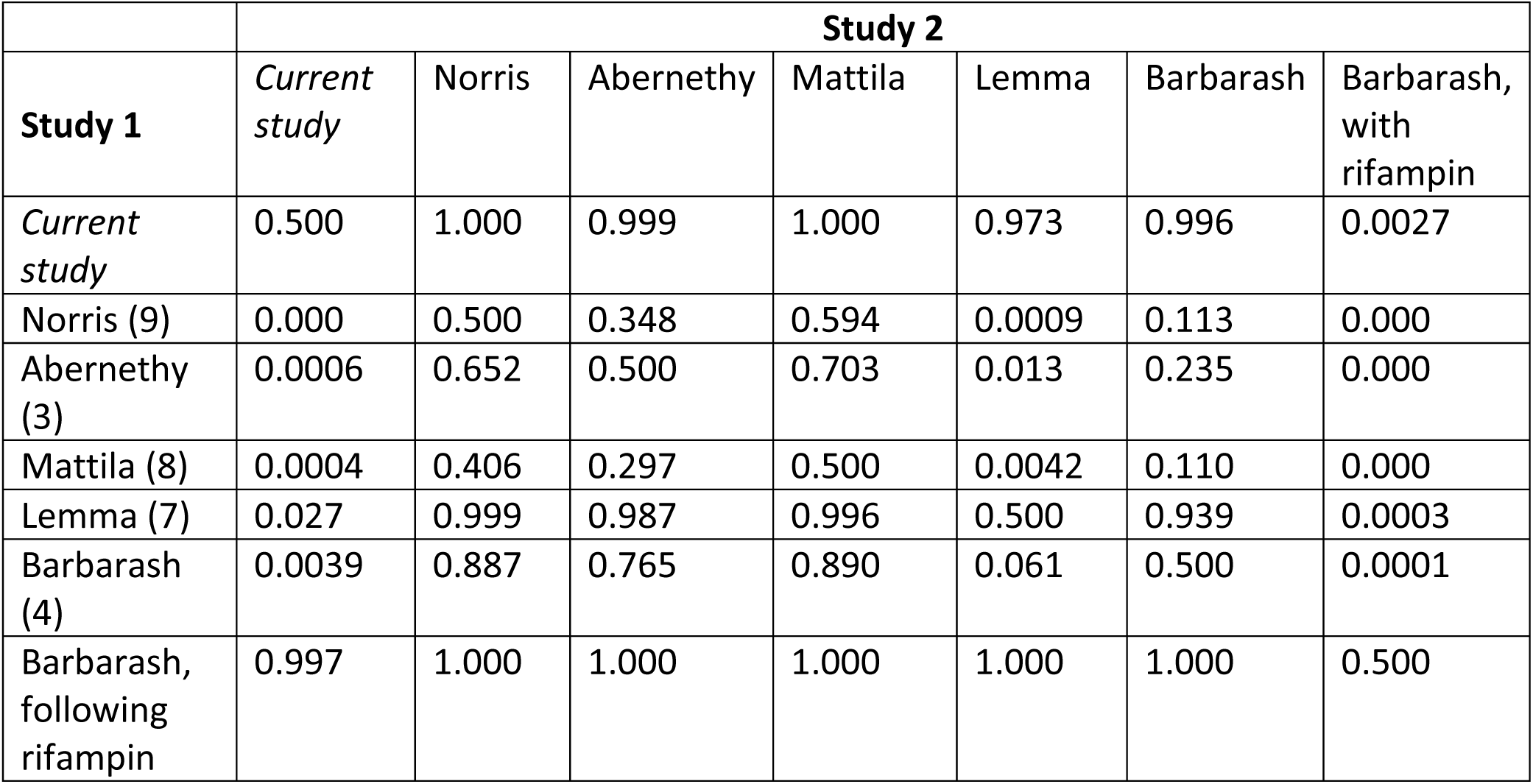
Probability table - Norverapamil to Verapamil AUC ratio versus historical controls (Supplement to Figure 3A) A probability greater than 0.95 indicates that the geometric mean norverapamil to verapamil AUC ratio in study 1 is significantly higher than that in study 2. The inverse situation, with probability <0.05, indicates that the geometric mean norverapamil to verapamil AUC ratio in study 1 is significantly lower than that in study 2.

|  | **Study 2** |  |  |  |  |  |  |
| --- | --- | --- | --- | --- | --- | --- | --- |
| **Study 1** | *Current study* | Norris | Abernethy | Mattila | Lemma | Barbarash | Barbarash, with rifampin |
| *Current study* | 0.500 | 1.000 | 0.999 | 1.000 | 0.973 | 0.996 | 0.0027 |
| Norris (9) | 0.000 | 0.500 | 0.348 | 0.594 | 0.0009 | 0.113 | 0.000 |
| Abernethy (3) | 0.0006 | 0.652 | 0.500 | 0.703 | 0.013 | 0.235 | 0.000 |
| Mattila (8) | 0.0004 | 0.406 | 0.297 | 0.500 | 0.0042 | 0.110 | 0.000 |
| Lemma (7) | 0.027 | 0.999 | 0.987 | 0.996 | 0.500 | 0.939 | 0.0003 |
| Barbarash (4) | 0.0039 | 0.887 | 0.765 | 0.890 | 0.061 | 0.500 | 0.0001 |
| Barbarash, following rifampin | 0.997 | 1.000 | 1.000 | 1.000 | 1.000 | 1.000 | 0.500 |

**Table S4c:** Probability table - R-verapamil to S-verapamil AUC ratio versus historical controls (Supplement to Figure 3B) A probability greater than 0.95 indicates that the geometric mean R-verapamil to S-verapamil AUC ratio in study 1 is significantly higher than that in study 2. The inverse situation, with probability <0.05, indicates that the geometric mean R-verapamil to S-verapamil AUC ratio in study 1 is significantly lower than that in study 2.

|  | **Study 2** |  |  |  |
| --- | --- | --- | --- | --- |
| **Study 1** | *Current study* | Lemma | Fromm | Fromm, with rifampin |
| *Current study* | 0.500 | 1.000 | 0.999 | 1.000 |
| Lemma (7) | 0.0001 | 0.500 | 0.0047 | 0.656 |
| Fromm (5) | 0.0006 | 0.995 | 0.50 | 0.999 |
| Fromm, with rifampin | 0.000 | 0.344 | 0.0014 | 0.5 |

**Table S4d:**
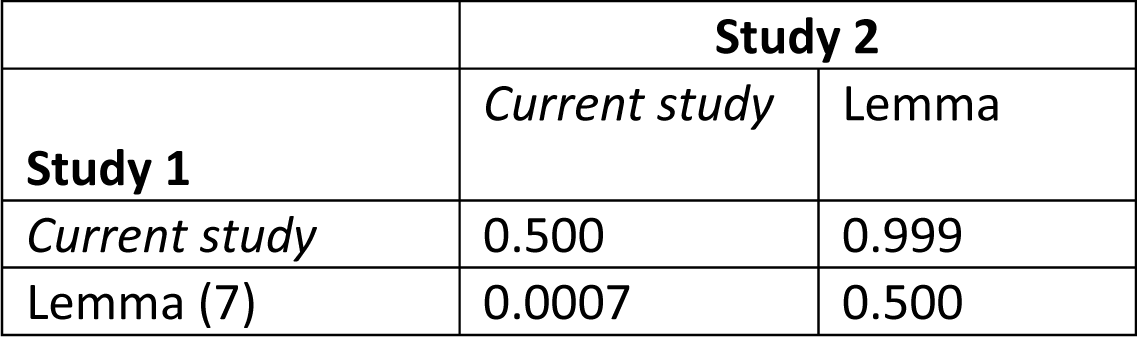
Probability table - R-norverapamil to S-norverapamil AUC ratio versus historical controls (Supplement to Figure 3C) A probability greater than 0.95 indicates that the geometric mean R-norverapamil to S-norverapamil AUC ratio in study 1 is significantly higher than that in study 2. The inverse situation, with probability <0.05, indicates that the geometric mean R-norverapamil to S-norverapamil AUC ratio in study 1 is significantly lower than that in study 2.

|  | **Study 2** |  |
| --- | --- | --- |
| **Study 1** | *Current study* | Lemma |
| *Current study* | 0.500 | 0.999 |
| Lemma (7) | 0.0007 | 0.500 |

**Table S4e:** Probability table - Verapamil AUC by genotype (Supplement to Figure 5) Probability greater than 0.95 indicates the restricted geometric mean verapamil AUC for genotype 1 is significantly higher than that of genotype 2. The inverse situation, with probability <0.05, indicates that the restricted geometric mean verapamil AUC of genotype 1 is significantly lower than that of genotype 2.

| SNP |  |  |  |
| --- | --- | --- | --- |
| **rs1045642 (ABCB1)** | **Genotype 2** |  |  |
| **Genotype 1** | **CC** | **TC** | **TT** |
| **CC** | 0.500 | **0.978** | **0.984** |
| **TC** | **0.022** | 0.500 | 0.665 |
| **TT** | **0.016** | 0.335 | 0.500 |
| **rs2032582 (ABCB1)** | **Genotype 2** |  |  |
| **Genotype 1** | **GG** | **TG** | **TT** |
| **GG** | 0.500 | 0.555 | 0.623 |
| **TG** | 0.445 | 0.500 | 0.598 |
| **TT** | 0.378 | 0.402 | 0.500 |
| **rs776746 (CYP3A5)** | **Genotype 2** |  |  |
| **Genotype 1** | **AA** | **AG** | **GG** |
| **AA** | 0.500 | 0.238 | 0.438 |
| **AG** | 0.762 | 0.500 | 0.711 |
| **GG** | 0.562 | 0.289 | 0.500 |

**Table S5.** Bioanalytical Validation Summary (SITEC Laboratory):

| Sr. No. | Experiment | Parameter | Summary of Results | Acceptance Criteria |
| --- | --- | --- | --- | --- |
| 1 | Solution Linearity | Correlation coefficient | 0.9997 | > 0.95 |
| 2 | Sensitivity | Signal to Noise ratio at LLOQ concentration | 123.7 | > 5.0 |
| 3 | Specificity | Interference in blank plasma | Six sources of plasma analyzed and met the acceptance criteria | < 20% of peak height at LLOQ concentration |
| 4 | Calibration curve | Concentration range | 9.9 to 1491.8 ng/mL | NA |
| 5 | Quality controls | Concentration | LLOQ QC: 9.9 ng/mL<br>LQC: 29.7 ng/mL<br>MQC-A: 198.0 ng/mL<br>MQC-B: 791.9 ng/mL<br>HQC: 1187.9 ng/mL | NA |
| 6 | Within batch precision | % CV at LLOQ QC | 1.13% to 3.00% | < 20% |
|  |  | % CV at LQC | 0.48% to 3.39% | < 15% |
|  |  | % CV at MQC - A | 0.46% to 2.09% | < 15% |
|  |  | % CV at MQC - B | 0.78% to 1.55% | < 15% |
|  |  | % CV at HQC | 0.70% to 2.21% | < 15% |
| 7 | Within batch accuracy | % Accuracy at LLOQ QC | 87.88% to 95.96% | 80.00% – 120.00% |
|  |  | % Accuracy at LQC | 94.28% to 103.70% | 85.00% - 115.00% |
|  |  | % Accuracy at MQC - A | 95.51% to 102.22% | 85.00% - 115.00% |
|  |  | % Accuracy at MQC - B | 92.78% to 96.98% | 85.00% - 115.00% |
|  |  | % Accuracy at HQC | 89.88% to 93.06% | 85.00% - 115.00% |
| 8 | Between batch precision | % CV at LLOQ QC | 3.72% | < 20% |
|  |  | % CV at LQC | 4.14% | < 15% |
|  |  | % CV at MQC - A | 2.91% | < 15% |
|  |  | % CV at MQC - B | 2.19% | < 15% |
|  |  | % CV at HQC | 1.90% | < 15% |
| 9 | Between batch accuracy | % Accuracy at LLOQ QC | 91.92% | 80.00% – 120.00% |
|  |  | % Accuracy at LQC | 97.64% | 85.00% - 115.00% |
|  |  | % Accuracy at MQC - A | 99.34% | 85.00% - 115.00% |
|  |  | % Accuracy at MQC - B | 94.08% | 85.00% - 115.00% |
|  |  | % Accuracy at HQC | 91.50% | 85.00% - 115.00% |
| 10 | Recovery | Mean % Recovery and %CV | % Recovery: 100.83<br>%CV range: 2.67 – 5.95 | %CV < 15% |
| 11 | Stock solution stability | % Stability | 103.63% @ 28 days | 85.00% - 115.00% |
| 12 | Short-term stability in plasma | % Stability at LQC & HQC | LQC: 98.46% @ 5 hrs<br>HQC: 87.46 @ 5 hrs | 85.00% - 115.00% |
| 13 | Freeze-thaw stability | % Stability at LQC & HQC after 3 cycles | LQC: 94.95%<br>HQC: 93.11% | 85.00% - 115.00% |
| 14 | Post-preparative stability @ room temperature | % Stability at LQC & HQC for 120 hrs | LQC: 104.38%<br>HQC: 94.07% | 85.00% - 115.00% |
| 15 | Dilution integrity | % Accuracy | Dilution factor 5: 91.95%<br>Dilution factor 3: 93.82% | 85.00% - 115.00% |

**S-verapamil:**
| Sr. No. | Experiment | Parameter | Summary of Results | Acceptance Criteria |
| --- | --- | --- | --- | --- |
| 1 | Solution Linearity | Correlation coefficient | 0.9994 | > 0.95 |
| 2 | Sensitivity | Signal to Noise ratio at LLOQ concentration | 20.8 | > 5.0 |
| 3 | Specificity | Interference in blank plasma | Six sources of plasma analyzed and met the acceptance criteria | < 20% of peak height at LLOQ concentration |
| 4 | Calibration curve | Concentration range | 2.0 ng/mL to 399.6 ng/mL | NA |
| 5 | Quality controls | Concentration | LLOQ QC: 2.0 ng/mL<br>LQC: 6.0 ng/mL<br>MQC-A: 50.1 ng/mL<br>MQC-B: 200.4 ng/mL<br>HQC: 320.6 ng/mL | NA |
| 6 | Within batch precision | % CV at LLOQ QC | 1.80% to 5.42% | < 20% |
|  |  | % CV at LQC | 1.33% to 3.17% | < 15% |
|  |  | % CV at MQC - A | 0.56% to 2.39% | < 15% |
|  |  | % CV at MQC - B | 0.92% to 1.69% | < 15% |
|  |  | % CV at HQC | 1.04% to 2.78% | < 15% |
| 7 | Within batch accuracy | % Accuracy at LLOQ QC | 90.00% to 105.00% | 80.00% – 120.00% |
|  |  | % Accuracy at LQC | 93.33% to 105.00% | 85.00% - 115.00% |
|  |  | % Accuracy at MQC - A | 95.21% to 105.19% | 85.00% - 115.00% |
|  |  | % Accuracy at MQC - B | 96.71% to 100.60% | 85.00% - 115.00% |
|  |  | % Accuracy at HQC | 96.38% to 98.32% | 85.00% - 115.00% |
| 8 | Between batch precision | % CV at LLOQ QC | 7.77% | < 20% |
|  |  | % CV at LQC | 4.74% | < 15% |
|  |  | % CV at MQC - A | 4.13% | < 15% |
|  |  | % CV at MQC - B | 2.02% | < 15% |
|  |  | % CV at HQC | 1.79% | < 15% |
| 9 | Between batch accuracy | % Accuracy at LLOQ QC | 100.00% | 80.00% – 120.00% |
|  |  | % Accuracy at LQC | 100.00% | 85.00% - 115.00% |
|  |  | % Accuracy at MQC - A | 100.40% | 85.00% - 115.00% |
|  |  | % Accuracy at MQC - B | 98.45% | 85.00% - 115.00% |
|  |  | % Accuracy at HQC | 97.66% | 85.00% - 115.00% |
| 10 | Recovery | Mean % Recovery and %CV | % Recovery: 107.49<br>%CV range: 3.10 – 5.09 | %CV < 15% |
| 11 | Stock solution stability | % Stability | 103.63% @ 28 days | 85.00% - 115.00% |
| 12 | Short-term stability in plasma | % Stability at LQC & HQC | LQC: 96.67 @ 5 hrs<br>HQC: 92.26 @ 5 hrs | 85.00% - 115.00% |
| 13 | Freeze-thaw stability | % Stability at LQC & HQC after 3 cycles | LQC: 101.67%<br>HQC: 97.79% | 85.00% - 115.00% |
| 14 | Post-preparative stability @ room temperature | % Stability at LQC & HQC for 120 hrs | LQC: 111.67%<br>HQC: 100.03% | 85.00% - 115.00% |
| 15 | Dilution integrity | % Accuracy | Dilution factor 5: 98.35%<br>Dilution factor 3: 96.69% | 85.00% - 115.00% |

**R-norverapamil:**
| Sr. No. | Experiment | Parameter | Summary of Results | Acceptance Criteria |
| --- | --- | --- | --- | --- |
| 1 | Solution Linearity | Correlation coefficient | 0.9996 | > 0.95 |
| 2 | Sensitivity | Signal to Noise ratio at LLOQ concentration | 88.9 | > 5.0 |
| 3 | Specificity | Interference in blank plasma | Six sources of plasma analyzed and met the acceptance criteria | < 20% of peak height at LLOQ concentration |
| 4 | Calibration curve | Concentration range | 6.0 ng/mL to 1001.6 ng/mL | NA |
| 5 | Quality controls | Concentration | LLOQ QC: 6.0 ng/mL<br>LQC: 18.0 ng/mL<br>MQC-A: 99.7 ng/mL<br>MQC-B: 498.6 ng/mL<br>HQC: 797.8 ng/mL | NA |
| 6 | Within batch precision | % CV at LLOQ QC | 1.30% to 1.79% | < 20% |
|  |  | % CV at LQC | 0.69% to 4.36% | < 15% |
|  |  | % CV at MQC - A | 0.54% to 1.93% | < 15% |
|  |  | % CV at MQC - B | 0.64% to 1.64% | < 15% |
|  |  | % CV at HQC | 0.69% to 2.62% | < 15% |
| 7 | Within batch accuracy | % Accuracy at LLOQ QC | 100.00% to 105.00% | 80.00% – 120.00% |
|  |  | % Accuracy at LQC | 103.33% to 109.44% | 85.00% - 115.00% |
|  |  | % Accuracy at MQC - A | 100.20% to 107.92% | 85.00% - 115.00% |
|  |  | % Accuracy at MQC - B | 99.30 to 102.77% | 85.00% - 115.00% |
|  |  | % Accuracy at HQC | 97.73% to 101.83% | 85.00% - 115.00% |
| 8 | Between batch precision | % CV at LLOQ QC | 1.30% | < 20% |
|  |  | % CV at LQC | 0.69% | < 15% |
|  |  | % CV at MQC - A | 0.81% | < 15% |
|  |  | % CV at MQC - B | 0.64% | < 15% |
|  |  | % CV at HQC | 0.69% | < 15% |
| 9 | Between batch accuracy | % Accuracy at LLOQ QC | 103.33% | 80.00% – 120.00% |
|  |  | % Accuracy at LQC | 105.56% | 85.00% - 115.00% |
|  |  | % Accuracy at MQC - A | 104.11% | 85.00% - 115.00% |
|  |  | % Accuracy at MQC - B | 101.08% | 85.00% - 115.00% |
|  |  | % Accuracy at HQC | 99.40% | 85.00% - 115.00% |
| 10 | Recovery | Mean % Recovery and %CV | % Recovery: 99.77<br>%CV range: 3.16 – 3.79 | %CV < 15% |
| 11 | Stock solution stability | % Stability | 99.83% @ 28 days | 85.00% - 115.00% |
| 12 | Short-term stability in plasma | % Stability at LQC & HQC | LQC: 101.11% @ 5 hrs<br>HQC: 97.27 @ 5 hrs | 85.00% - 115.00% |
| 13 | Freeze-thaw stability | % Stability at LQC & HQC after 3 cycles | LQC: 100.56%<br>HQC: 104.80% | 85.00% - 115.00% |
| 14 | Post-preparative stability @ room temperature | % Stability at LQC & HQC for 120 hrs | LQC: 109.44%<br>HQC: 97.99% | 85.00% - 115.00% |
| 15 | Dilution integrity | % Accuracy | Dilution factor 5: 98.70%<br>Dilution factor 3: 96.85% | 85.00% - 115.00% |

**S-norverapamil:**
| Sr. No. | Experiment | Parameter | Summary of Results | Acceptance Criteria |
| --- | --- | --- | --- | --- |
| 1 | Solution Linearity | Correlation coefficient | 0.9994 | > 0.95 |
| 2 | Sensitivity | Signal to Noise ratio at LLOQ concentration | 28.6 | > 5.0 |
| 3 | Specificity | Interference in blank plasma | Six sources of plasma analyzed and met the acceptance criteria | < 20% of peak height at LLOQ concentration |
| 4 | Calibration curve | Concentration range | 2.0 to 394.5 ng/mL | NA |
| 5 | Quality controls | Concentration | LLOQ QC: 2.0 ng/mL<br>LQC: 6.0 ng/mL<br>MQC-A: 49.9 ng/mL<br>MQC-B: 199.6 ng/mL<br>HQC: 319.4 ng/mL | NA |
| 6 | Within batch precision | % CV at LLOQ QC | 2.44% to 6.27% | < 20% |
|  |  | % CV at LQC | 0.86% to 5.42% | < 15% |
|  |  | % CV at MQC - A | 0.73% to 2.16% | < 15% |
|  |  | % CV at MQC - B | 0.48% to 2.08% | < 15% |
|  |  | % CV at HQC | 0.63% to 1.66% | < 15% |
| 7 | Within batch accuracy | % Accuracy at LLOQ QC | 90.00% to 100.00% | 80.00% – 120.00% |
|  |  | % Accuracy at LQC | 98.33% to 103.33% | 85.00% - 115.00% |
|  |  | % Accuracy at MQC - A | 97.60% to 106.21% | 85.00% - 115.00% |
|  |  | % Accuracy at MQC - B | 96.74% to 105.41% | 85.00% - 115.00% |
|  |  | % Accuracy at HQC | 96.06% to 105.95% | 85.00% - 115.00% |
| 8 | Between batch precision | % CV at LLOQ QC | 6.46% | < 20% |
|  |  | % CV at LQC | 3.72% | < 15% |
|  |  | % CV at MQC - A | 3.46% | < 15% |
|  |  | % CV at MQC - B | 3.70% | < 15% |
|  |  | % CV at HQC | 4.05% | < 15% |
| 9 | Between batch accuracy | % Accuracy at LLOQ QC | 95.00% | 80.00% – 120.00% |
|  |  | % Accuracy at LQC | 101.67% | 85.00% - 115.00% |
|  |  | % Accuracy at MQC - A | 101.40% | 85.00% - 115.00% |
|  |  | % Accuracy at MQC - B | 100.25% | 85.00% - 115.00% |
|  |  | % Accuracy at HQC | 100.03% | 85.00% - 115.00% |
| 10 | Recovery | Mean % Recovery and %CV | % Recovery: 98.82<br>%CV range:0.87 – 4.39 | %CV < 15% |
| 11 | Stock solution stability | % Stability | 103.60% @ 28 days | 85.00% - 115.00% |
| 12 | Short-term stability in plasma | % Stability at LQC & HQC | LQC: 101.67% @ 5 hrs<br>HQC: 97.40 @ 5 hrs | 85.00% - 115.00% |
| 13 | Freeze-thaw stability | % Stability at LQC & HQC after 3 cycles | LQC: 100.00%<br>HQC: 99.97% | 85.00% - 115.00% |
| 14 | Post-preparative stability @ room temperature | % Stability at LQC & HQC for 120 hrs | LQC: 103.33%<br>HQC: 94.02% | 85.00% - 115.00% |
| 15 | Dilution integrity | % Accuracy | Dilution factor 5: 97.56%<br>Dilution factor 3: 95.18% | 85.00% - 115.00% |

**Figure S1A:**
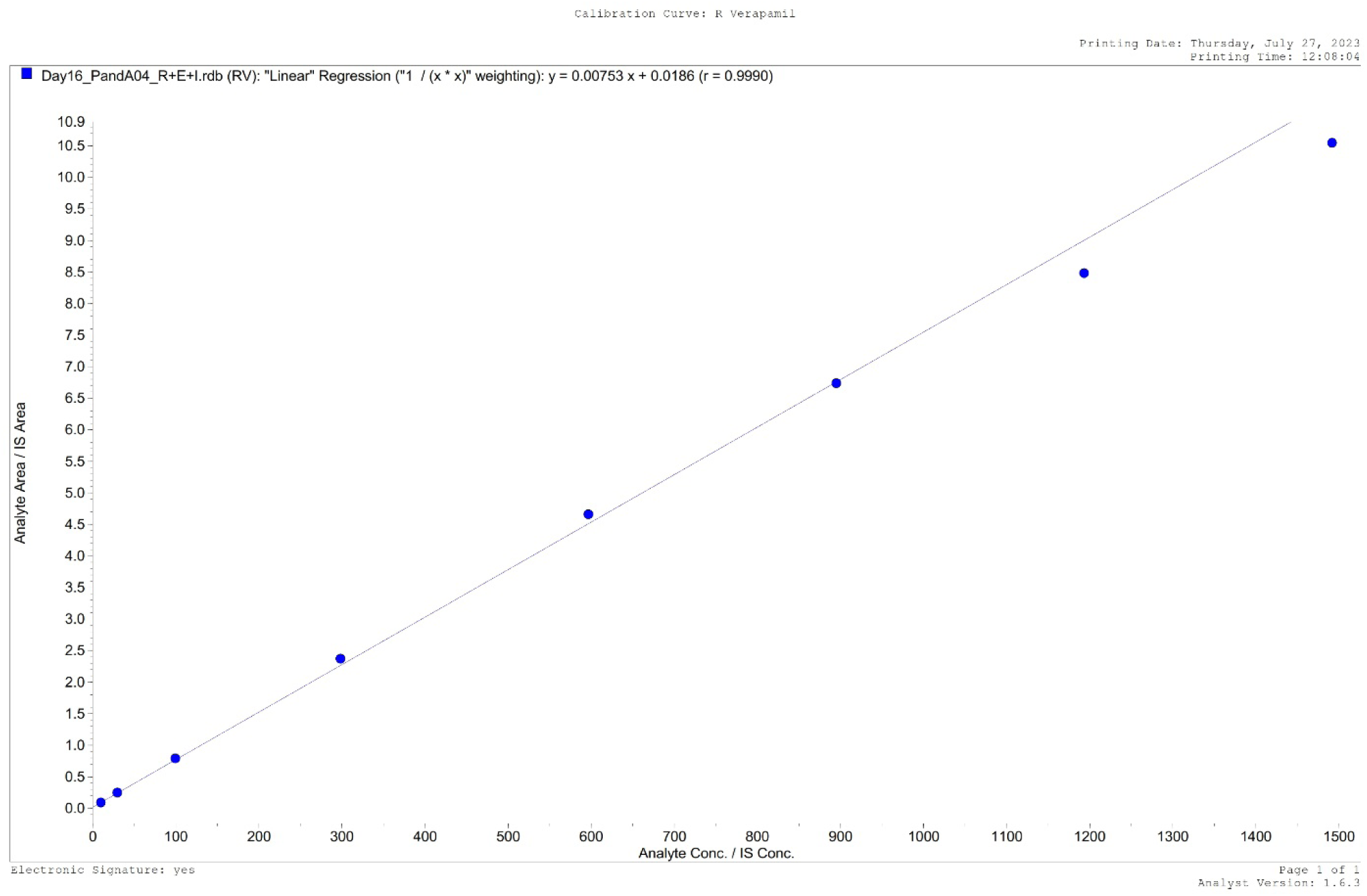
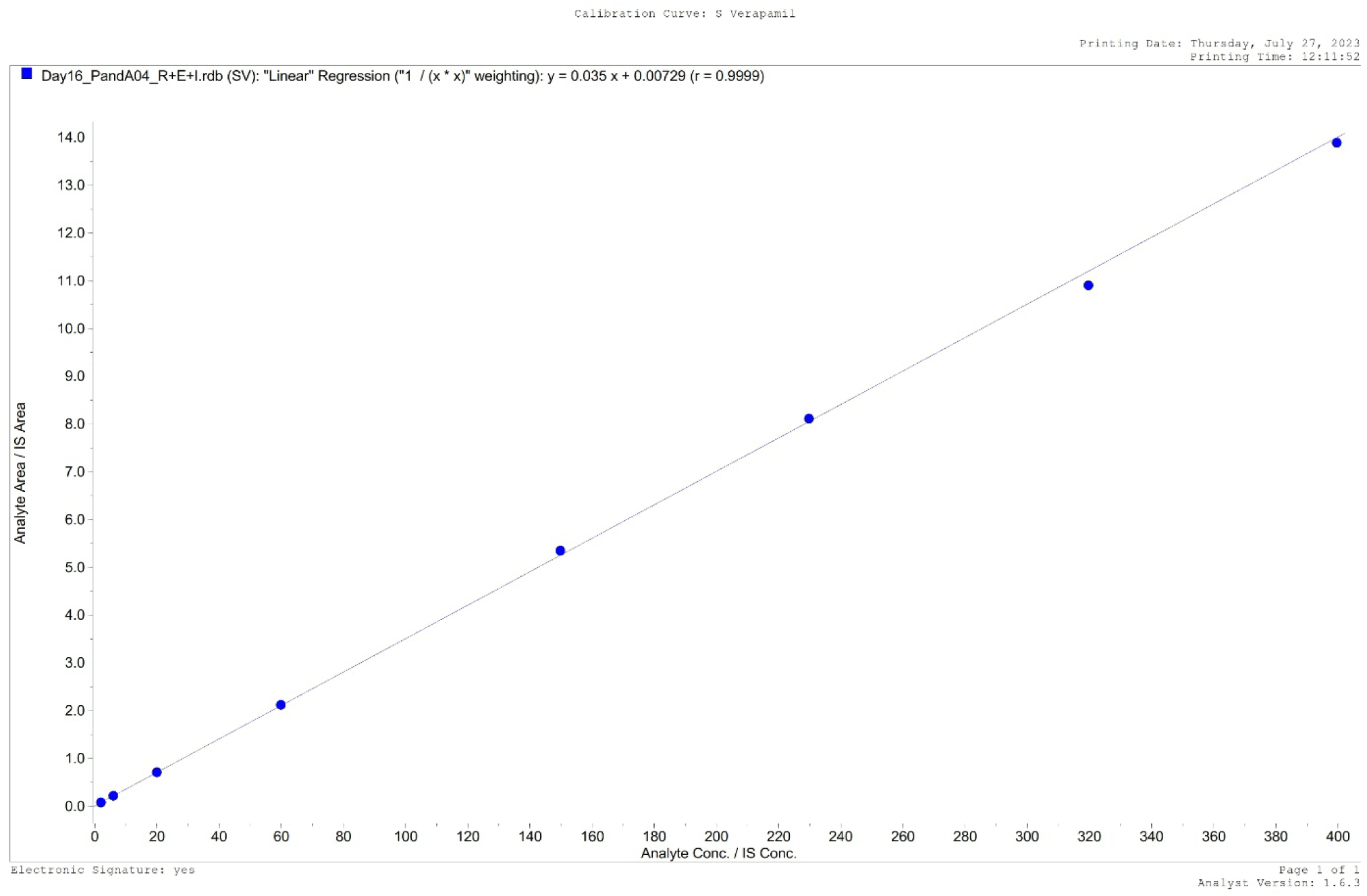
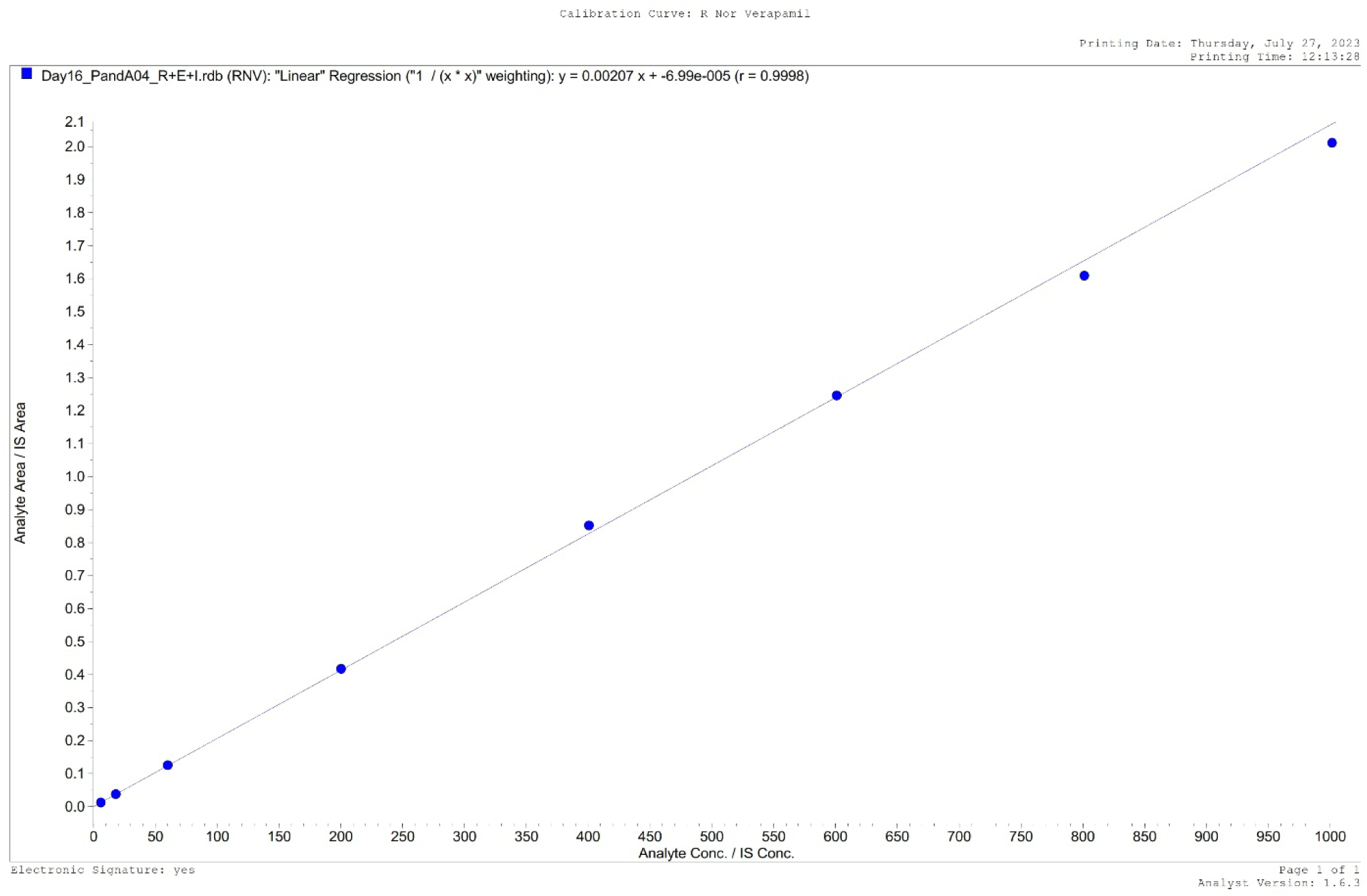
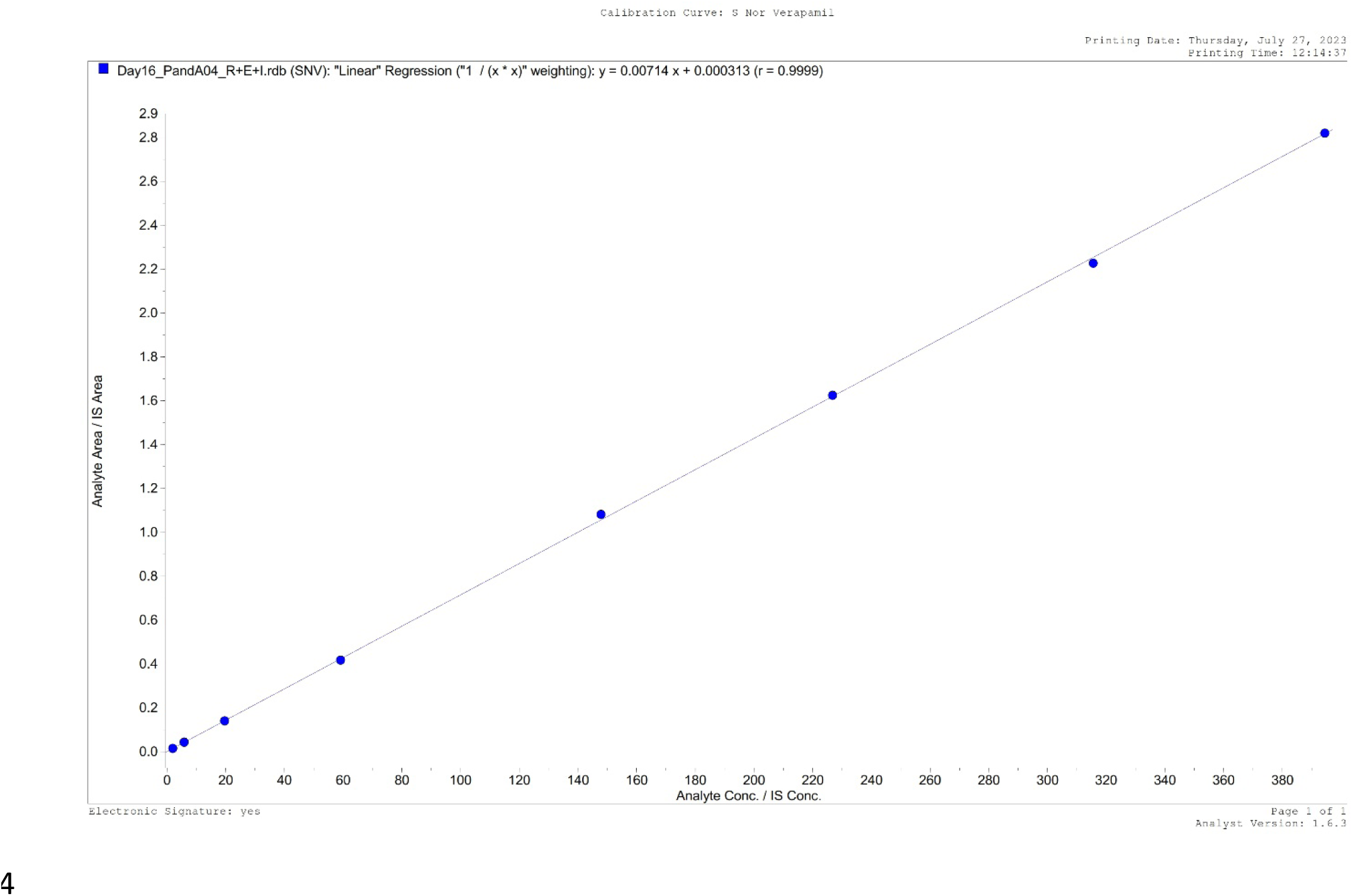
Individual participant measurements of verapamil, norverapamil and their enantiomers – see supplementary figure S1 file. Levels of verapamil (total, R-and S-enantiomers) and norverapamil (total, R- and S-enantiomers) obtained on study day 9 are shown for each participant. **Figure S1B: Individual participant measurement of verapamil and norverapamil by laboratory – see supplementary figure S1 file** Levels of verapamil and norverapamil obtained on study day 9 are shown for each study participant as measured by NIRT and SITEC laboratories respectively.

---

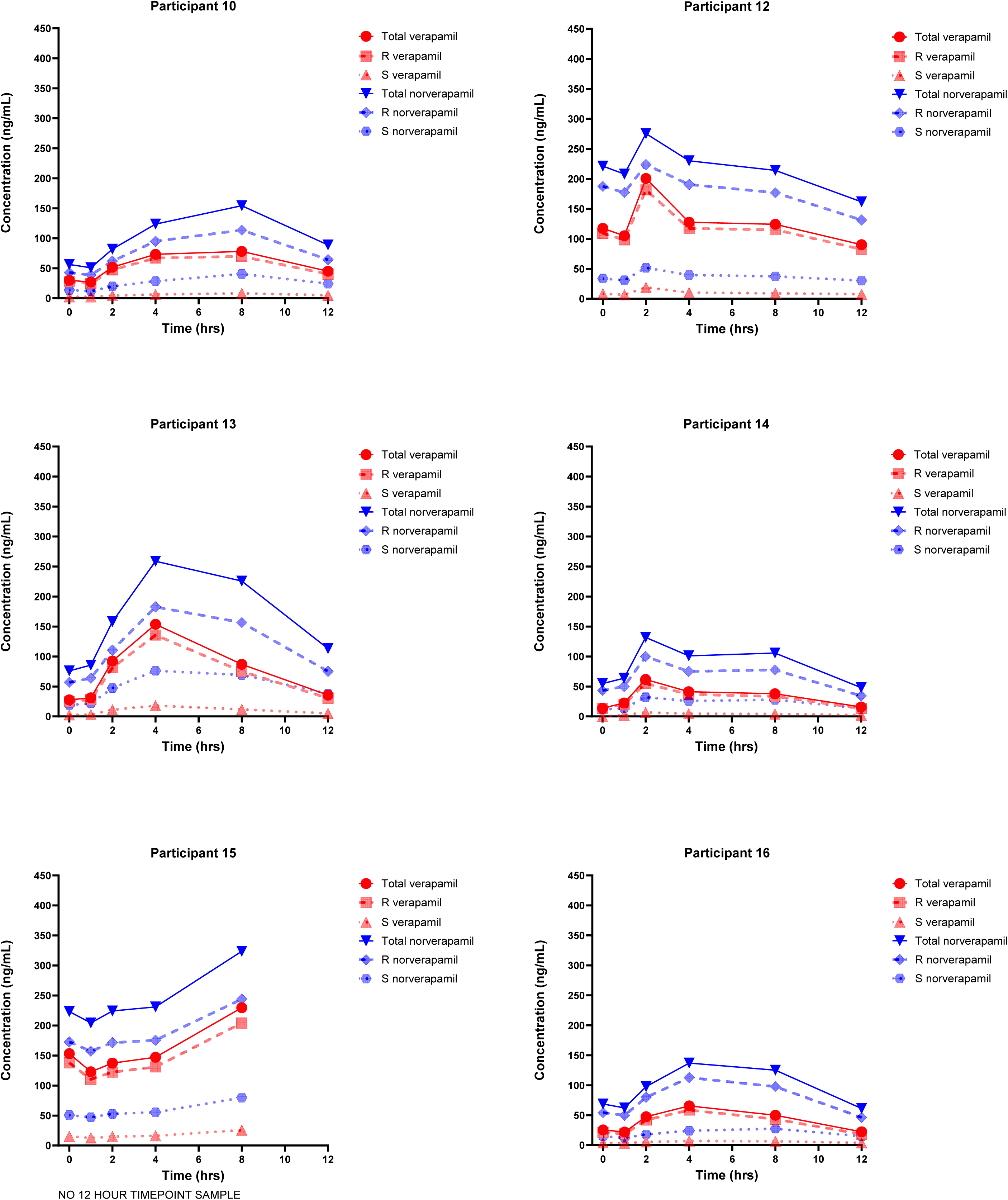

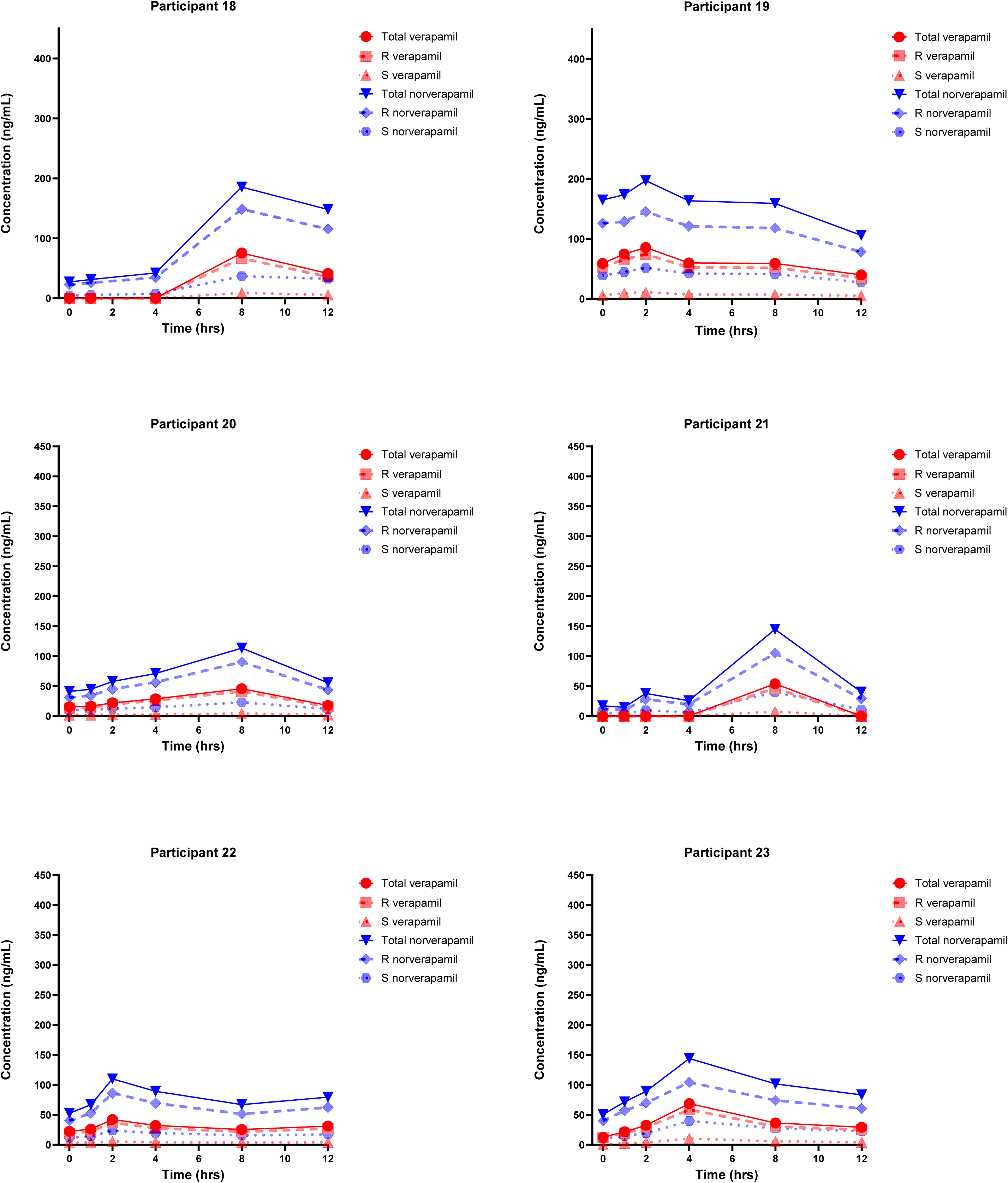

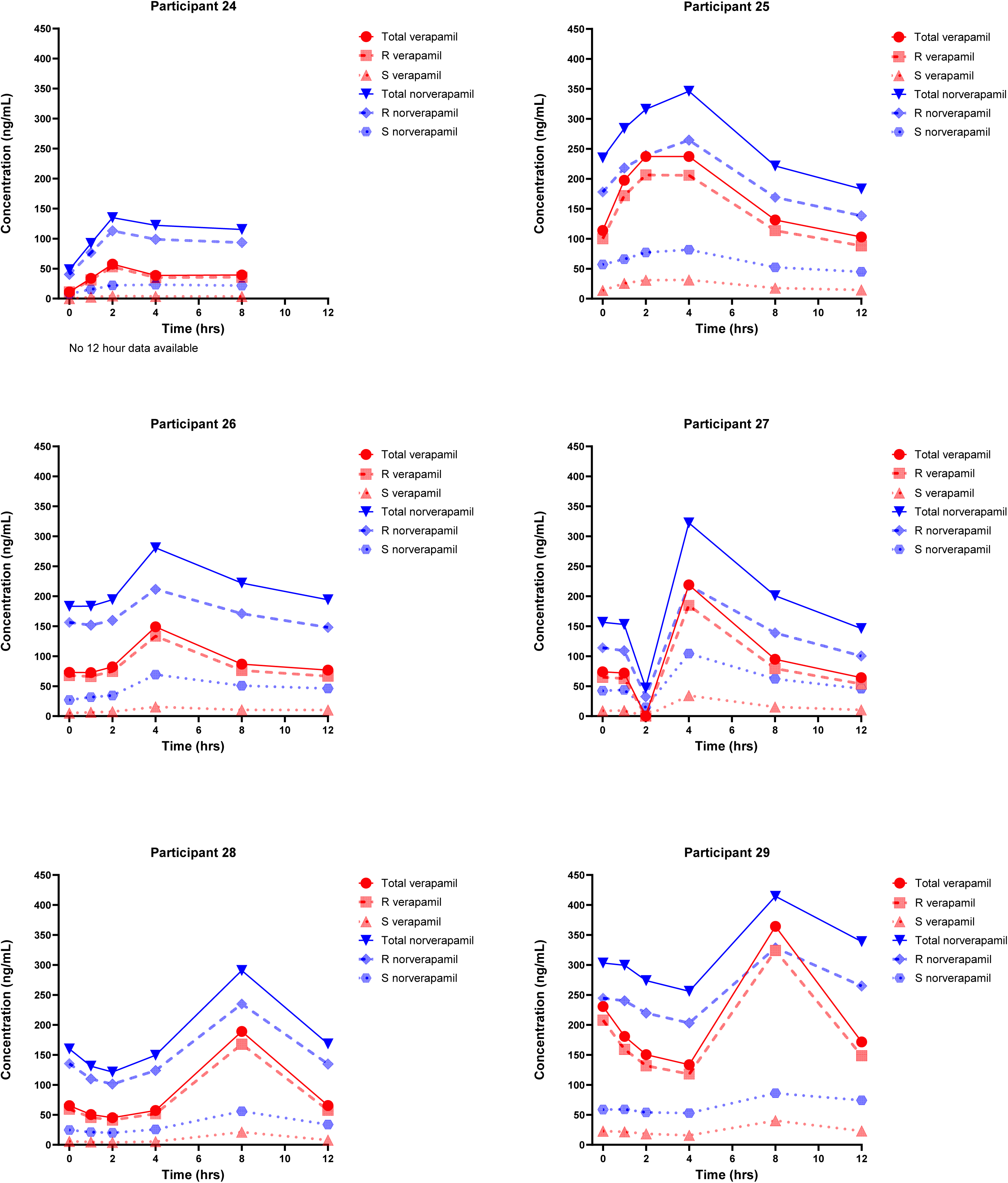

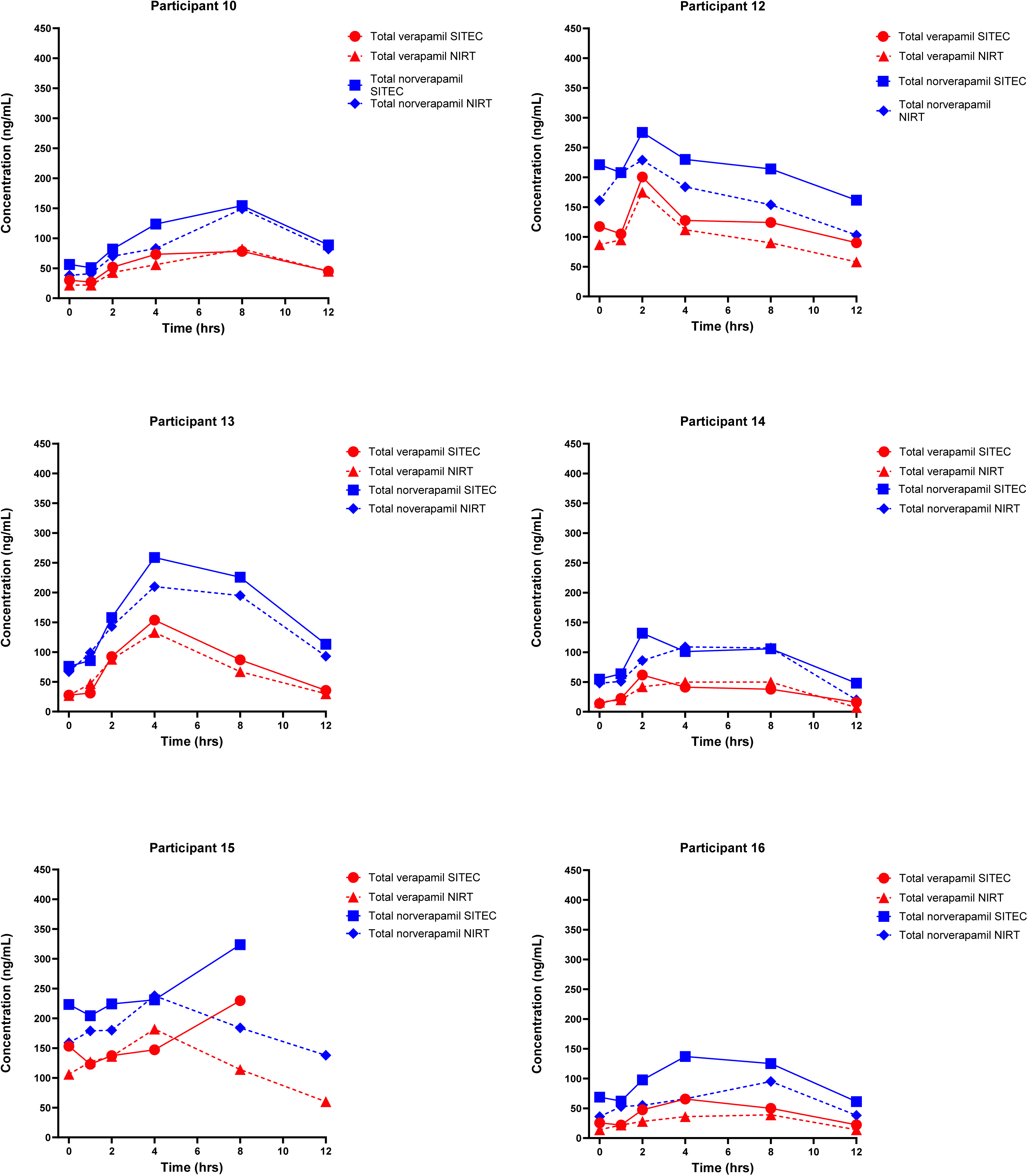

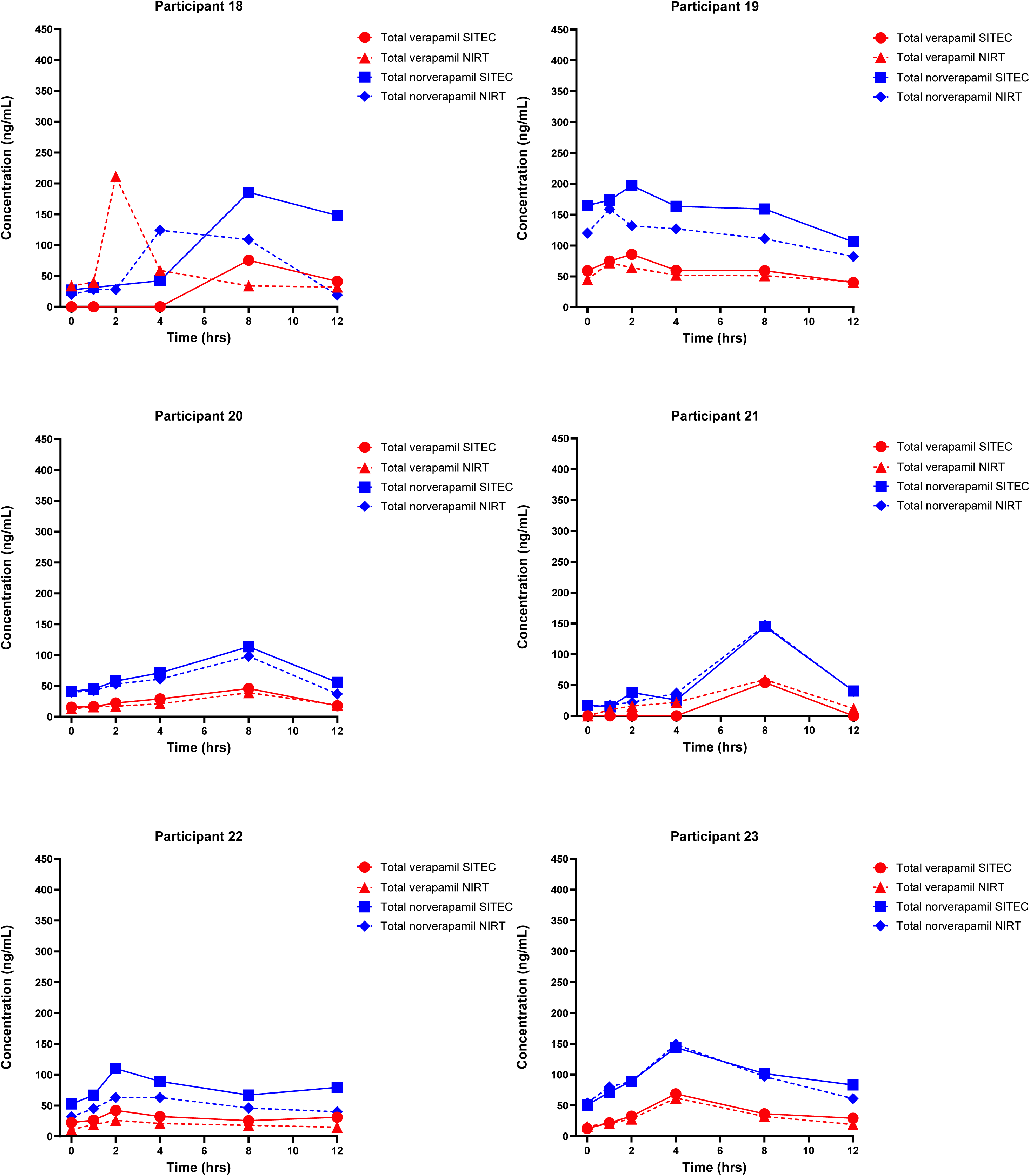

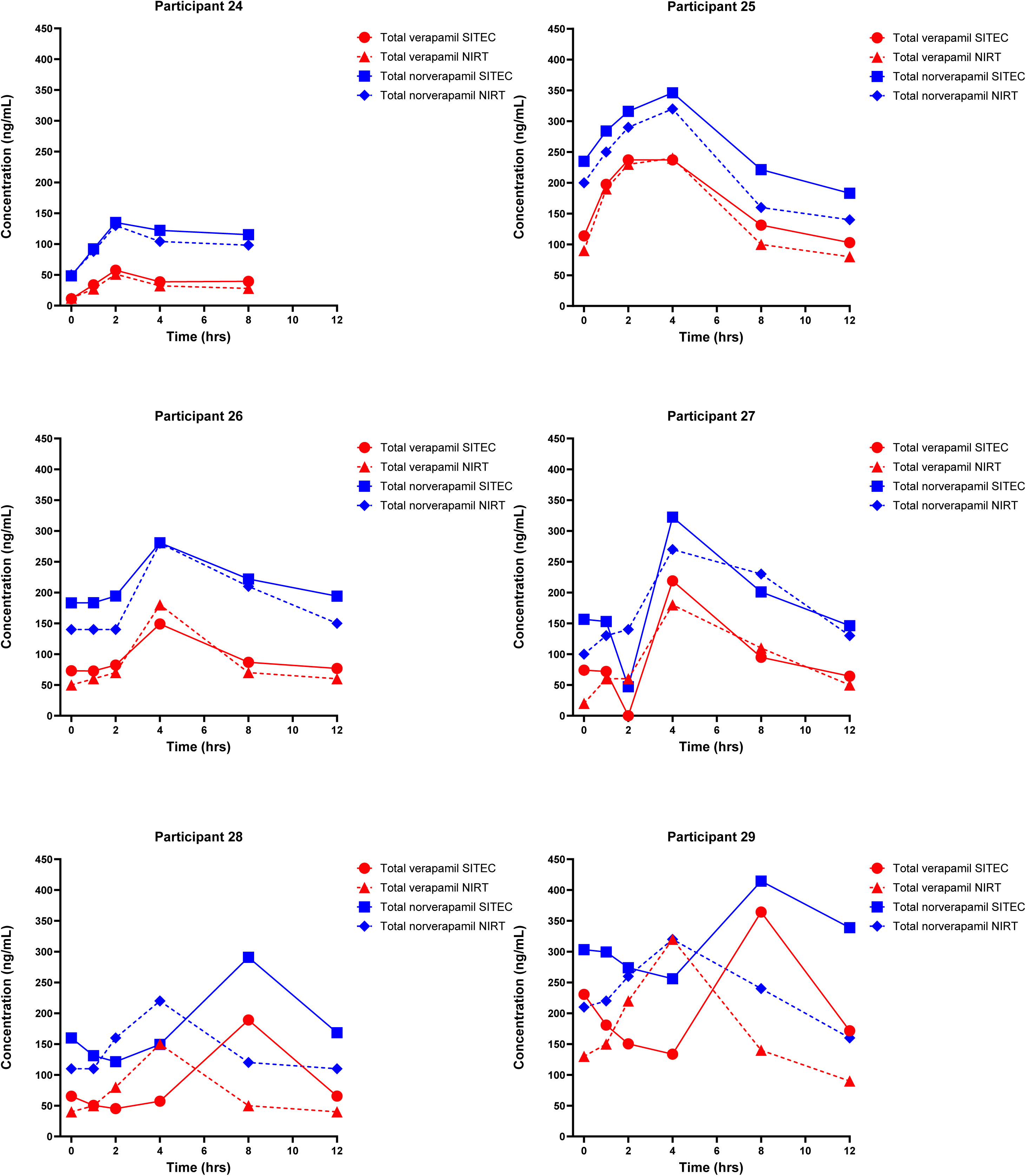

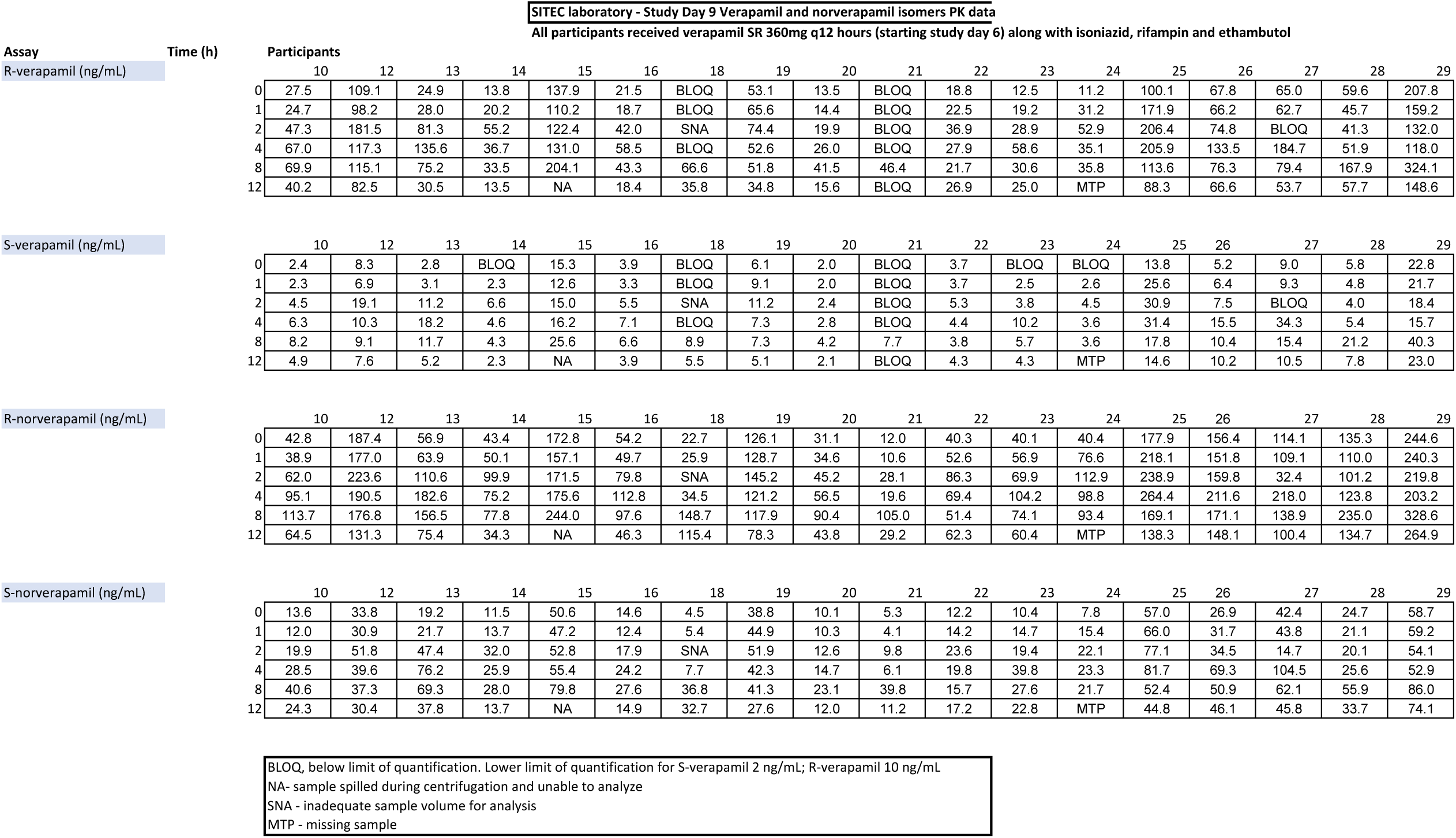

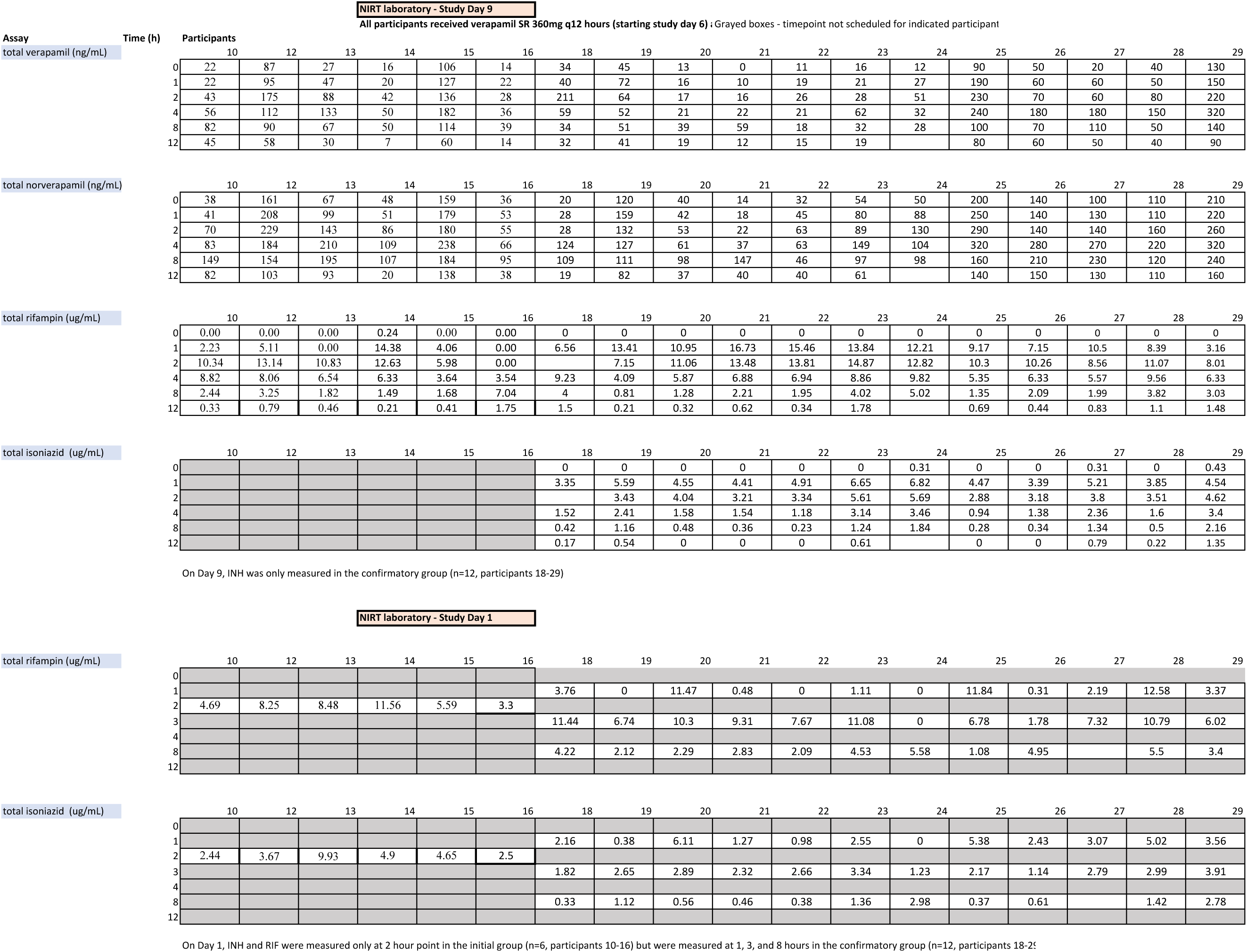

## Statistical Appendix 1 Bayesian Methods

### 1 Introduction – The five problems discussed

We adopt the Bayesian paradigm for addressing all these problems.

We first address the comparison of verapamil AUCs by SNP genotype.

Turning then to ratios of norverapamil to verapamil (or R- to S-verapamil, or R- to S-norverapamil), we then discuss how we determined to use the log-Gaussian (a.k.a log-normal) distribution to model all the distributions of verapamil, its metabolites, and their enantiomers other than for comparison by genotype.

Next we address how to set an appropriate prior on the parameters of the 2-dimensional log-Gaussian arising.

We then look at how we inferred the properties of the log-Gaussian distributions from which samples had been taken both in this study (for which the raw data was available) and in the various previous studies referenced in the main paper (for which the raw data was not available, only the sample means and sample standard deviations), and how to determine the properties of the resulting log-Gaussian distributions on the ratios of norverapamil to verapamil or R enantiomer to S enantiomer.

Finally we discuss how paired values of rifampicin or isoniazid AUCs pre- and post-verapamil were assessed.

### 2 Notation and units

Irrespective of notation elsewhere in this paper, in this supplementary appendix on methods we use the following notation except where stated otherwise:

*K* The number of patients

*N* The number of dimensions involved; this will be 2 where the input data is a pair of values (e.g. of S and R enantiomer) for each patient, or 1 where the input data is a single value (e.g. the ratio of R enantiomer to S enantiomer) for each patient.

Units for Area Under Curve (AUC) and derived quantities will be microgram-hours per litre throughout. Throughout, if *V* is a vector or matrix, then *e^V^* will denote the matrix *W* whose (*j, k*)-element *w_j,k_* is given by *e^vj,k^*, and not the matrix exponential 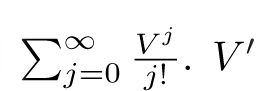 will denote the transpose of *V*.

### 3 The first problem – Comparison of AUCs between different genotypes at a particular SNP

Readers who have not encountered Bayesian methods before, and Bayesian model choice in particular, may like to read an informal intuitive introduction in Appendix 1 of [1] and details of Bayesian model choice for identifying distributions in Appendix 2 of [1]. In this section we adopt the notation of that Appendix 2, noting only the points at which the present analysis differs from the analysis of distribution type done there. All references to [1] are to that Appendix 2 unless stated otherwise.

Specifically we proceed exactly as in section 2 of [1], using the same range of distributions, but with some different prior parameters as discussed below. No decision is taken on which distribution type is present; rather the posterior probability that the restricted geometric mean of one genotype is greater than that of another is calculated using the basic rules of probability, integrating out the distribution type variable.

#### 3.1 Priors used

We use prior 1 from [1] section 4, i.e. the same probability of ^1^ on each of the distributions listed in section 3 of [1].

For the parameters of the individual distributions, the general comments in [1] section 5.1 apply, as do the specific parameters given in subsequent sections apart from the following:

##### 3.1.1 Gaussian

We take the values used in [1] section 5.2, except with *µ_µ_* = 1, *s_s_* = 0.04.

##### 3.1.2 Student

We take the values used in [1] section 5.4, except with *µ_µ_* = 1, *s_µ_* = 0.25, *s_s_* = 25.

##### 3.1.3 skew-Student

We take the values used in [1] section 5.8, except with *µ_ξ_* = 1, *s_ξ_* = 0.25, *s_s_* = 25.

### 4 The second problem – Identification of the distribution family for the purpose of calculations of ratios of AUCs

As in section 3, we adopt the methods of [1] Appendix 2, but in this case stopping after applying Bayesian model choice as discussed in part 1 of section 2 of [1] (again all references to [1] are to that paper’s Appendix 2).

We use the same priors as discussed in section 3.1 above.

#### 4.1 Results on distribution family

The posterior probabilities of the verapamil and norverapamil AUCs being distributed according to the various distribution families considered are shown in table 1, while the corresponding analysis for each enantiomer separately is shown in table 2.

Therefore, for this particular application it turns out that one particular distribution family, the log-Gaussian family, is both most probable and most convenient for subsequent calculation, given that the ratio of two log-Gaussian variables is also log-Gaussian (even if the logarithms of the two variables are jointly non-independently Gaussian, i.e. correlated, as in this case).

We therefore forego the the equivalent of part 2 of section 2 of [1] and will proceed to address our third problem under an assumption of log-Gaussianity.

**Table 1:** Posterior probabilities of the distribution of AUC values of verapamil, norverapamil, or both, being of the type shown. The entries in the column headed “Both” are the posterior probabilities given additional assumptions that the distributions of both verapamil and norverapamil AUCs must be from the same family, and that the posterior probabilities are independent.

|  | Total verapamil AUC | Total norverapamil AUC | Both |
| --- | --- | --- | --- |
| Gaussian | 0.0000 | 0.0042 | 0.0000 |
| logGaussian | 0.2409 | 0.2200 | 0.3163 |
| Student | 0.0001 | 0.0000 | 0.0000 |
| logStudent | 0.2084 | 0.1172 | 0.1452 |
| Gamma | 0.1365 | 0.2198 | 0.1782 |
| invGamma | 0.1539 | 0.2187 | 0.1999 |
| skewStudent | 0.0008 | 0.0002 | 0.0000 |
| logskewStudent | 0.1572 | 0.0849 | 0.0792 |
| Gammapow | 0.1021 | 0.1339 | 0.0812 |

**Table 2:** Posterior probabilities of the distribution of AUC values of the different enantiomers of verapamil and norverapamil, or all four, being of the type shown. The entries in the column headed “All” are the posterior probabilities given additional assumptions that the distributions of each enantiomer’s AUCs must be from the same family and that the posterior probabilities are independent.

|  | S-verap AUC | R-verap AUC | S-norver AUC | R-norver AUC | All four |
| --- | --- | --- | --- | --- | --- |
| Gaussian | 0.0000 | 0.0000 | 0.0058 | 0.0022 | 0.0000 |
| logGaussian | 0.2344 | 0.2409 | 0.2178 | 0.2239 | 0.5098 |
| Student | 0.0000 | 0.0000 | 0.0026 | 0.0015 | 0.0000 |
| logStudent | 0.2406 | 0.2072 | 0.1254 | 0.1192 | 0.1380 |
| Gamma | 0.1393 | 0.1350 | 0.2565 | 0.1906 | 0.1703 |
| invGamma | 0.1012 | 0.1585 | 0.1340 | 0.2601 | 0.1035 |
| skewStudent | 0.0005 | 0.0005 | 0.0021 | 0.0024 | 0.0000 |
| logskewStudent | 0.1747 | 0.1568 | 0.0920 | 0.0864 | 0.0403 |
| Gammapow | 0.1093 | 0.1011 | 0.1637 | 0.1138 | 0.0381 |

## 5 The third problem – how to determine an appropriate prior on the parameters of the log-Gaussian

For this we used the conjugate joint prior to (*µ, S*), the mean and scale (inverse of covariance) parameters of a 2-d Gaussian, given by

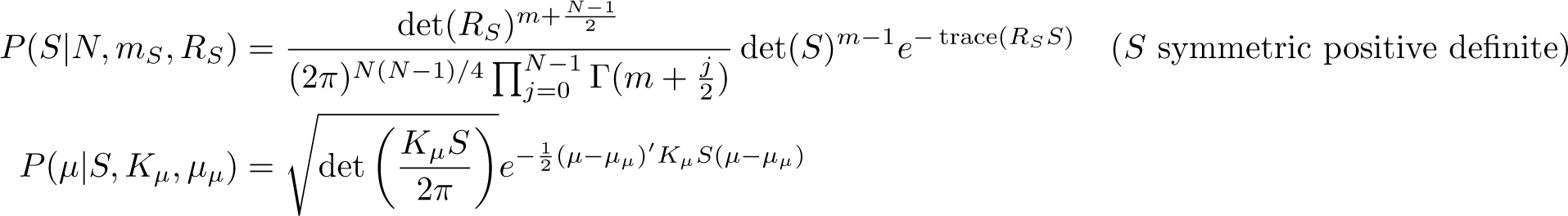

noting that *S* is then Wishart distributed on the *N* (*N* + 1)*/*2-dimensional submanifold of R*^N^*^2^ corresponding to symmetric positive definite matrices, rather than on the corresponding open subset of R*^N^*^(*N*+1)*/*2^. We initially set the very broad spherically symmetric (and in the *µ*-axis improper) priors

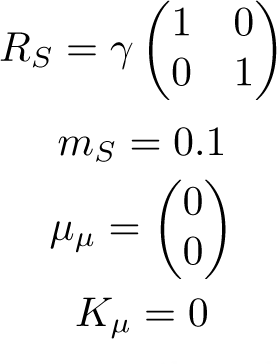

with *γ* = ^1^ for total verapamil : total norverapamil, and with *γ* = ^1^ for ratios of enantiomers. We then considered the data composed of the logarithms of raw AUC values for verapamil and norverapamil in the present study (16 points in all, each consisting of one value for verapamil AUC and one for norverapamil AUC). We calculated the resulting posterior distribution, giving corresponding new values of the parameters *m_S_, R_S_, µ_µ_, K_µ_*, and divided *m_S_* and *R_S_* by 10 (keeping the same mean scale matrix (and therefore same correlation at that mean) but broadening the prior), kept the same value of *µ_µ_*, and set *K_µ_* to 1. The resulting parameters were

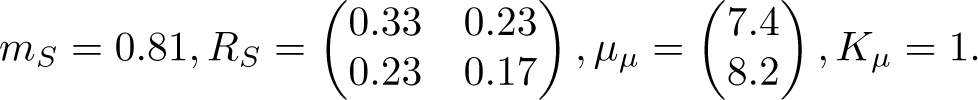

For the other cases, namely S- and R-verapamil, and S- and R-norverapamil, we followed the same procedure, getting the parameter sets

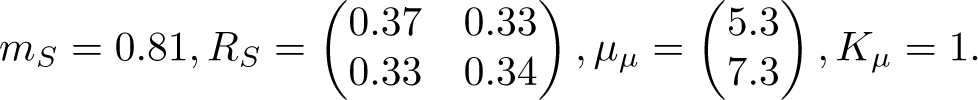

and

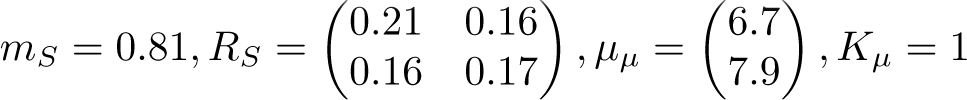

respectively.

## 6 The fourth problem – How to compare two distributions for which you have different types of data

### 6.1 Overview

Now, in this particular situation we want to compare the distributions of the ratios of AUCs for (e.g.) verapamil and norverapamil in individual patients with similar quantities occurring in previous studies.

However, in the present study we have raw data consisting of each individual’s AUC for verapamil and that for norverapamil, while in the previous studies, despite best efforts to obtain raw data from their authors, we have only the mean and standard deviation of the sample of AUCs for verapamil and for norverapamil – or in some cases, only the mean and standard deviation of the ratios.

In order to infer the mean and centiles of the distribution of the ratios in the population, we apply the standard techniques of Bayesian inference (referenced in [1]). Assuming, for the reasons given in section 4 above, that the AUCs are log-Gaussianly distributed across the population, and noting that the distribution of the 2 *×* 1 vector *v* consisting of an individual’s log verapamil AUC and his log norverapamil AUC is not spherically Gaussian but has positive correlation, we proceed specifically as follows.

Let *µ, S* be the unknown mean and scale-matrix parameters of the distribution of *v*. Let *z* denote the 2 *× K* matrix of *K* values of *e^v^*, and *x* the matrix of *K* values of *v*. The values of *m_S_, S_S_, µ_µ_*, and *K_µ_* given in section 5 above determine a prior probability distribution on *µ* and *S*. We know the distribution of *x* given *µ* and *S*, and we know the value of *z* = *e^x^* corresponding to any *x*, and for any *z* we know the values of the resulting

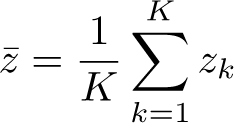

and

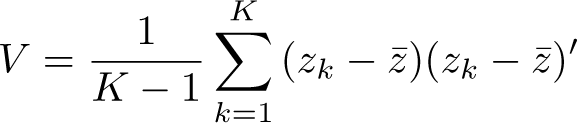

and

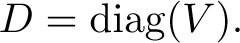

In the case that we have the value of *z* (i.e. the raw data) available, our next job is now to calculate *P* (*µ, S|z*), or more specifically to draw *L* (e.g. 10,000) random samples of (*µ, S*) from this distribution – method described in section 6.2 below. In the case that we do not know *z* but only know *z̄* and *D* we instead draw samples from the distribution *P* (*µ, S|z̄*, *D*). Either way we next calculate the distribution of the ratio *r* = *z*_1_*/z*_2_ for each such sample, which is again log-Gaussian with parameters

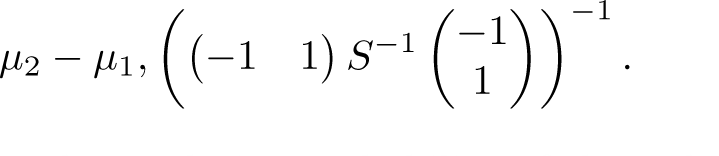

The usual formulae for the mean and cumulative probability of the 1-d log-Gaussian now allow us to determine, for each sample of (*µ, S*), the mean of the resulting distribution of the ratio *r* and any centiles we may be interested in. Accumulating e.g. the mean for each sample as a histogram, we can determine the posterior mean and centiles of distribution of the mean of *r* given whichever data it is that is available to us for this particular study, and similarly for the posterior mean and centiles of whichever centiles of the distribution of *r* we may be interested in.

Finally, to estimate the probability that e.g. the mean of *r* across the population in Study 1 is greater than that in Study 2, we use the *L* samples of the mean from each study to provide *L*^2^ comparisons, and count the fraction of such comparisons in which the mean of *r* in Study 1 is larger than in Study 2 as our estimate of the probability that the population mean of *r* in Study 1 is larger that that in Study 2.

### 6.2 Random sampling

The case that the raw data *z* is known is easy - we simply calculate the posterior distribution of (*µ, S*) and sample from it directly, getting a sequence of independent samples; sampling from the Wishart distribution is done using the Bartlett decomposition [2].

The case that *z* is unknown and that we have only observed *z̄* and *V* is harder. In this case we proceed by using Gibbs sampling (see [1]) to alternately resample *x* and (*µ, S*) given the other and (*z̄*, *D*). The case of resampling (*µ, S*) given *x* (and hence *z, z̄*, and *D*) is the same as the case where *z* is known.

To resample *x* given (*µ, S, z̄*, *D*) is harder. We note that this involves taking random samples from a distribution on R^2*K*^ given that the samples lie on an embedded curved (2*K −* 4)-dimensional manifold *M_z_*_̄,*D*_. The calculation of the probability density on the manifold is explained in [3]; the key fact is that the probability density given that *x ∈ M* is given by

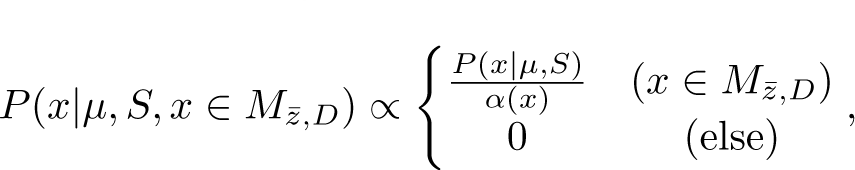

where

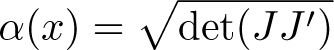

where

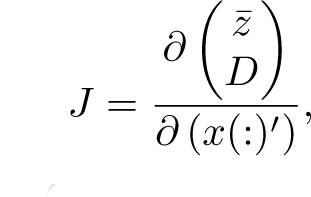

where *x*(:)*^I^* denotes the 1 *×* 2*K* row vector (*x*_1,1_, *x*_2,1_, *x*_1,2_, *…, x_K_*_,2_).

Specifically, we proceed using the Metropolis-Hastings algorithm [4]. We introduce as auxiliary variables a positive definite diagonal 2 *×* 2 matrix *A* and a 2 *×* 1 vector *y*, and put an arbitrary everywhere-supported probability distribution on the product space of such objects. Starting from *x*, we propose by first choosing a value of (*A, y*) at random from that distribution, then a value of *x*_prop_ from *P* (*x|µ, S*), then solving for the unique (*A*_new_, *y*_new_) such that *y*_new_ +*A*_new_*x*_prop_ *∈ M_z_*_̄,*D*_ by any suitable method, e.g. Newton-Raphson in four dimensions [5]. Finally we accept *x*_new_ = *y*_new_ + *A*_new_*x*_prop_ with probability

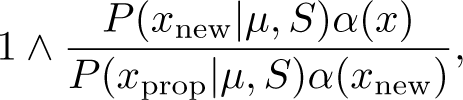

while otherwise we keep the old value of *x* (and discard (*A, y*) and (*A*_new_, *y*_new_) either way). Detailed balance holds by reversal of this exact path in (*x, A, y*) space and integrating over all such paths.

### 6.3 Implementation and testing

We implemented the above method in Matlab®. We tested the software using synthetic data drawn from the priors, in all three potential use cases, namely: (a) with the simulated raw data available; (b) when provided only with the values of *z̄* and *V* ; and (c) when provided only with the values of the mean and standard deviation of the ratios across the data sample.

With a number of data points up to 20, convergence of the Markov chain Monte-Carlo process was essentially immediate, with acceptance rates between 0.15 and 0.5. The near-immediate convergence (in the case where the raw data was not available) is illustrated by plots of the mean ratio over the population, the low centile of the ratio, and the coefficients of the correlation matrix of *x*, in figures 1, 2, 3 respectively.

We note that this method would need modification with large numbers of data points in order to improve the acceptance rate, for example by only updating a random subset of them at a time conditional on the those not being updated as well as on the other relevant variables. As a result convergence would be slower and potentially more samples would be needed. In the present case we took 10,000 samples for each analysis, keeping all but the first 1,000, which as can be seen from the figures is easily sufficient.

**Figure 1:**
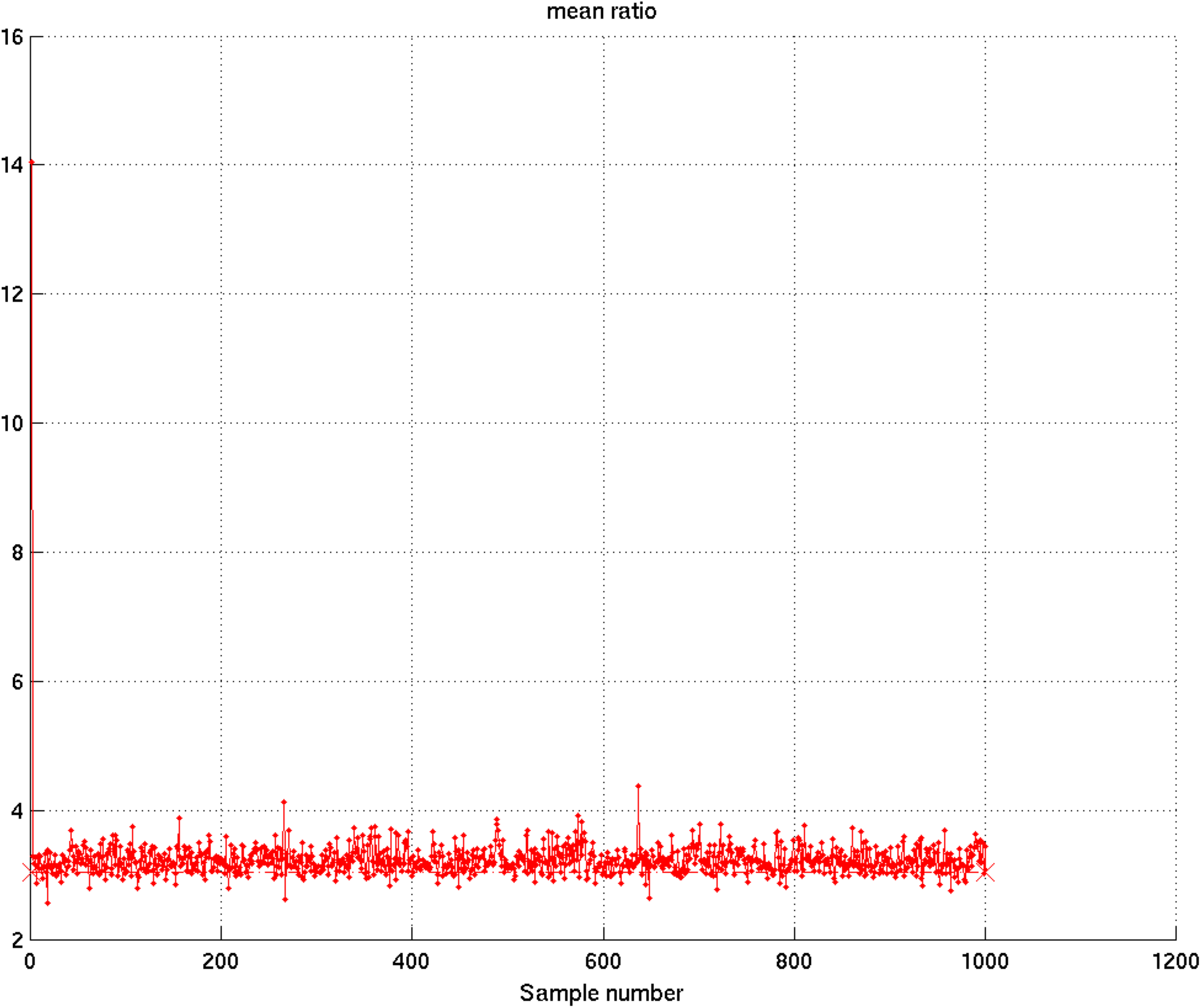
The sequence of samples of the mean of the ratio of the two substances over the population in a run using synthetic data. The true value is marked by a horizontal line with X markers at each end. It can be seen how convergence is essentially immediate after the first couple of samples, and how the distribution covers the true value.

**Figure 2:**
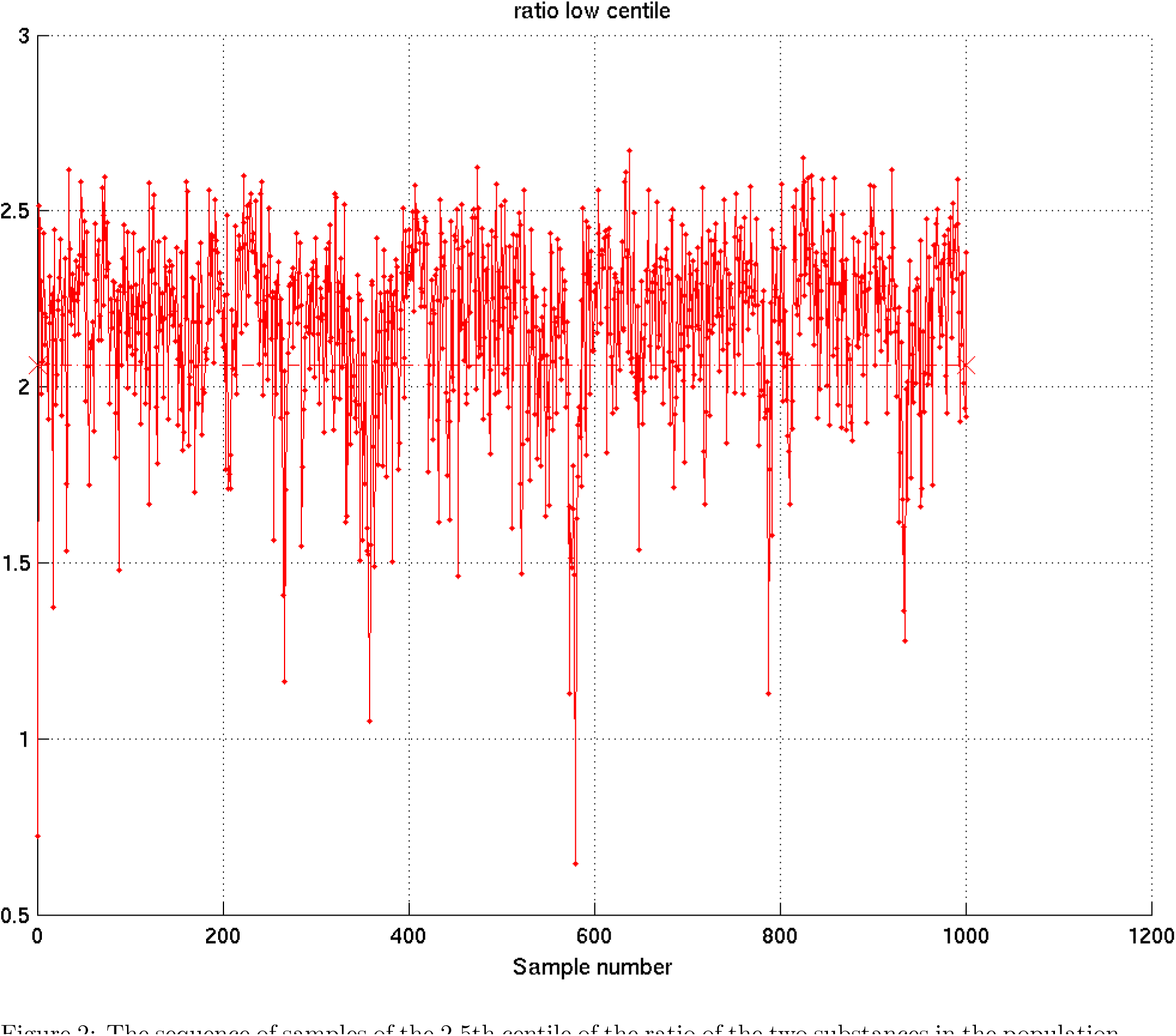
The sequence of samples of the 2.5th centile of the ratio of the two substances in the population in a run using synthetic data. The true value is marked by a horizontal line with X markers at each end. It can be seen how convergence is essentially immediate after the first couple of samples, and how the distribution covers the true value.

**Figure 3:**
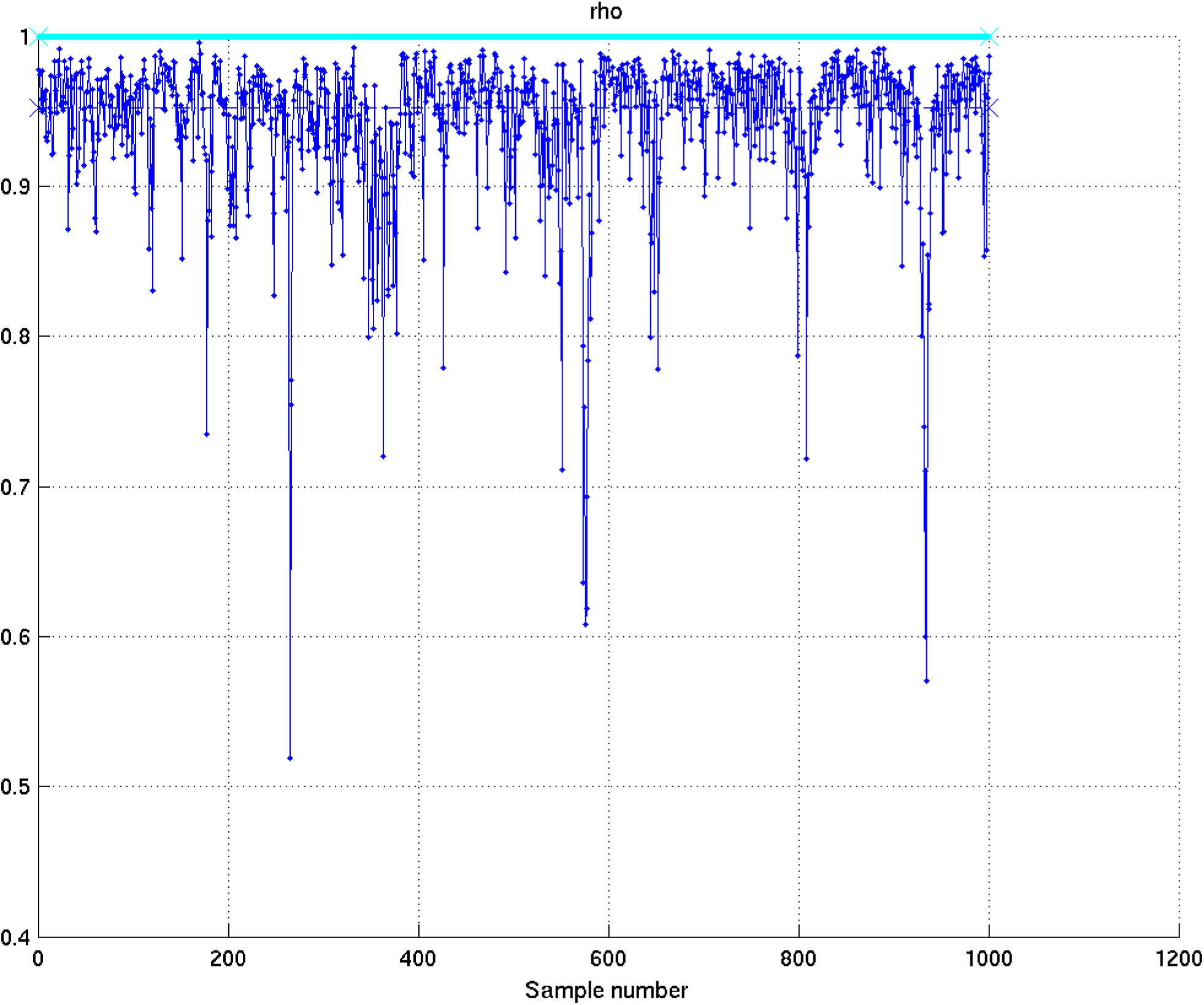
The sequence of samples of the correlation matrix of x in a run using synthetic data; the cyan (and red – covered by cyan) plots are of the (2, 2) (and (1, 1)) entries which are of course fixed at 1; the blue (and green – covered by blue) plots are of the (2, 1) (and (1, 2)) entries of the matrix, and represent the correlation coefficient of the values of the two substances across the population. The true value in each case is marked by a horizontal line with X markers at each end. It can be seen how convergence is essentially immediate after the first couple of samples, and how the distribution covers the true value, as well as how the prior permits correct modeling of the high level of correlation between the two substances.

### 6.4 Illustration of results on norverapamil/verapamil ratio

We illustrate the resulting information on the actual data in the present study with figures 4, 5, 6. In each case the green histogram comes from the actual observed data. Each blue curve represents a random sample from the posterior distribution of the distribution of the relevant quantity in the population. Clearly, given only 16 samples of (say) verapamil AUC, we cannot precisely determine what the distribution in the population is – so the blue curves illustrate what it might be, given the priors used and the data in question.

For each blue curve, we can determine its 2.5th centile (left-pointing triangle), its median (up-pointing triangle), its mean (cross), and its 97.5th centile (right-pointing triangle). Then, given several thousand such blue curves (not shown for obvious reasons), we can establish the mean and centiles of each of these properties of the distribution. The 2.5th and 97.5th centiles of each of these properties are shown at the top of the figure in red, illustrating the central 95% Bayesian confidence interval for that property of the distribution in the population.

In the case of previous studies, the green histogram is not available, so the samples of the distribution in the population come only from the priors used and the given mean and standard deviation of the samples of the relevant quantity. Getting them involves working out what the data samples might have been, and indeed considering many hypothetical sample values of the dataset in question. The entire process is mathematically rigorous and consistent with the laws of probability.

**Figure 4:**
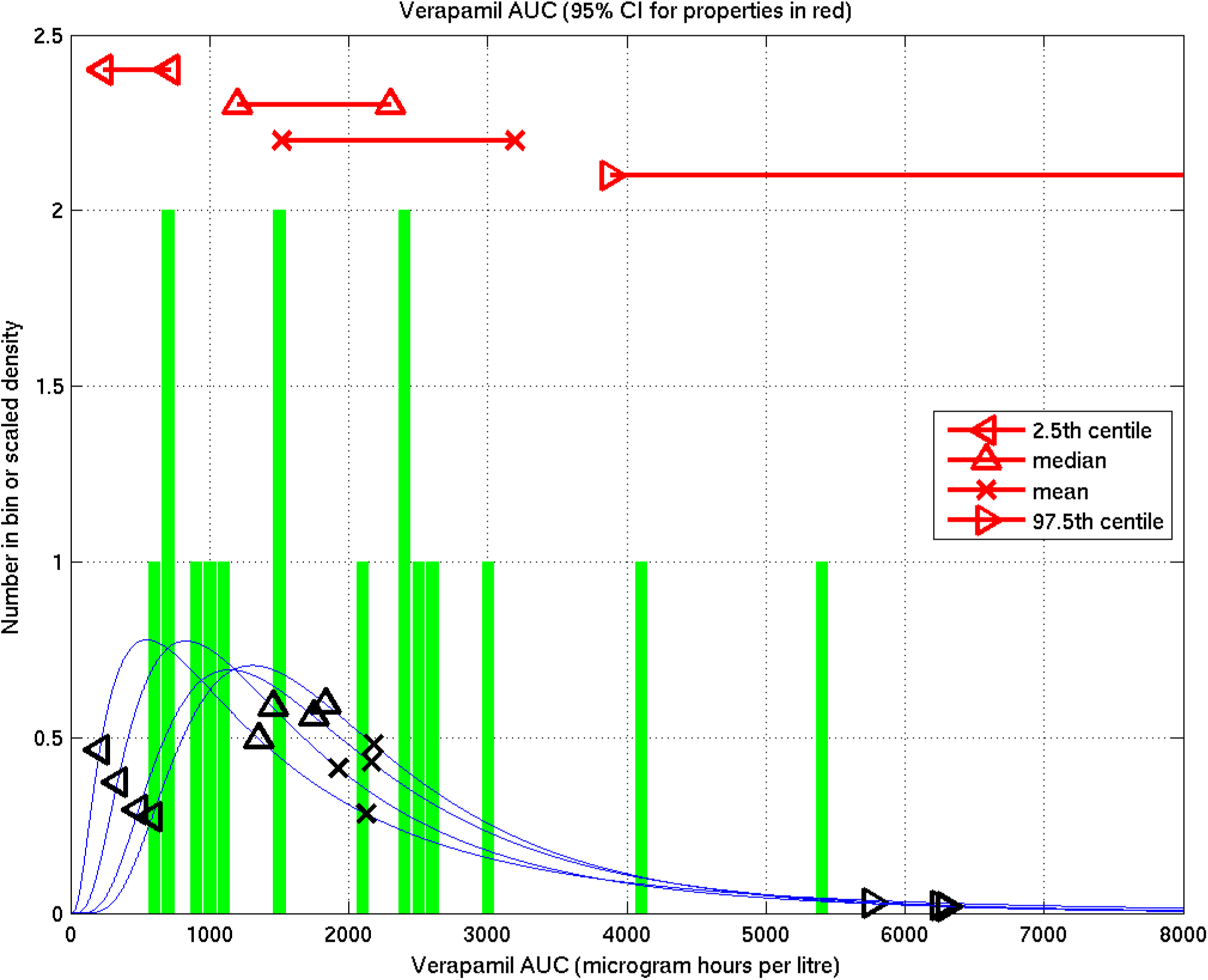
Inference from the present study on verapamil AUCs: The green histogram represents the actual measured values. Each blue curve represents a sample from the posterior distribution given that data of the distribution in the population. For each blue curve, the black left-pointing triangle is the 2.5th centile, the up-pointing triangle the median, the cross the mean, and the right-pointing triangle the 97.5th centile. In red at the top of the figure are the central 95% Bayesian confidence intervals for each of these quantities, based on 9000 samples of the distribution over the population (i.e. on 9000 such blue curves).

**Figure 5:**
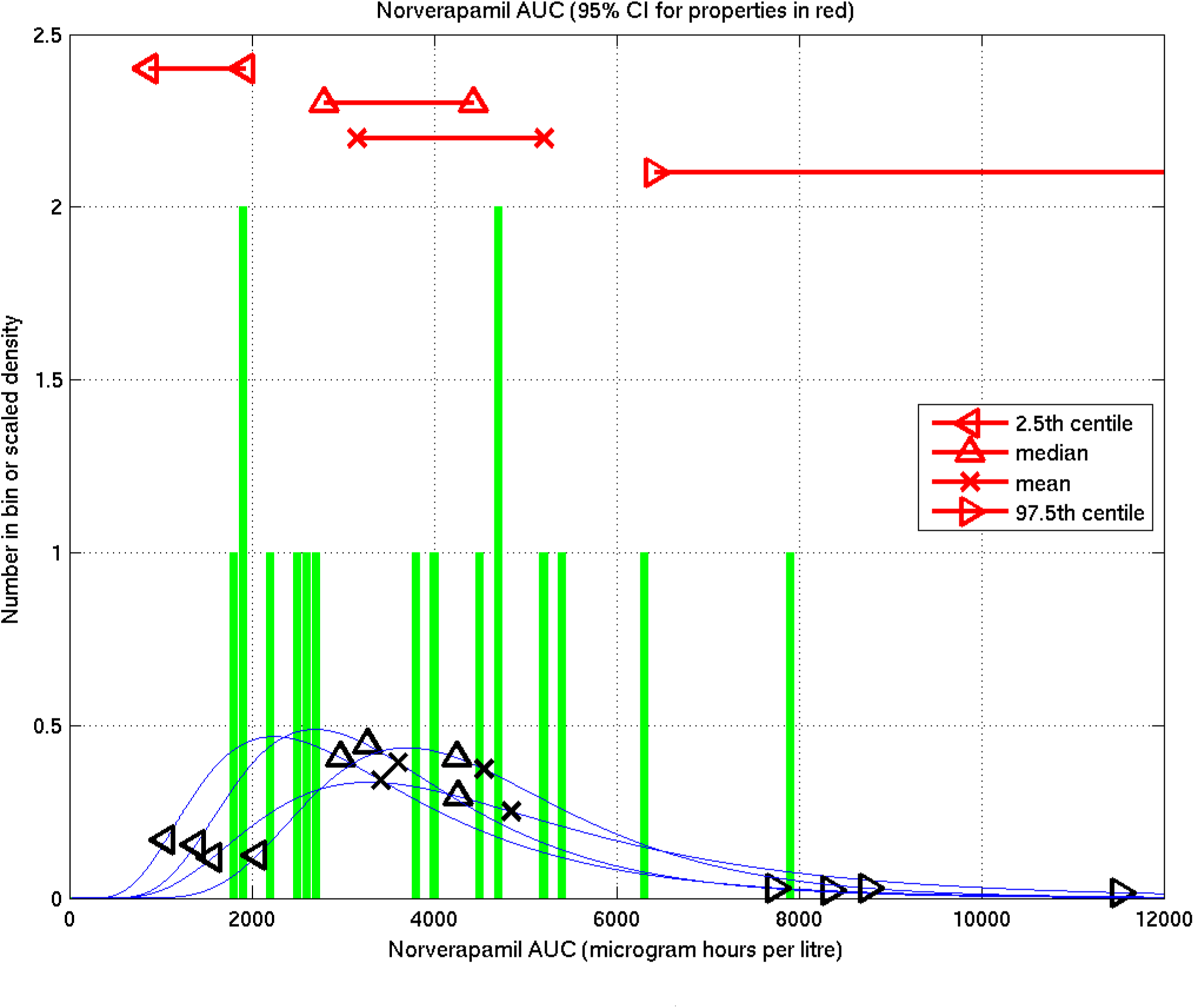
Inference from the present study on norverapamil AUCs: The green histogram represents the actual measured values. Each blue curve represents a sample from the posterior distribution given that data of the distribution in the population. For each blue curve, the black left-pointing triangle is the 2.5th centile, the up-pointing triangle the median, the cross the mean, and the right-pointing triangle the 97.5th centile. In red at the top of the figure are the central 95% Bayesian confidence intervals for each of these quantities, based on 9000 samples of the distribution over the population (i.e. on 9000 such blue curves).

**Figure 6:**
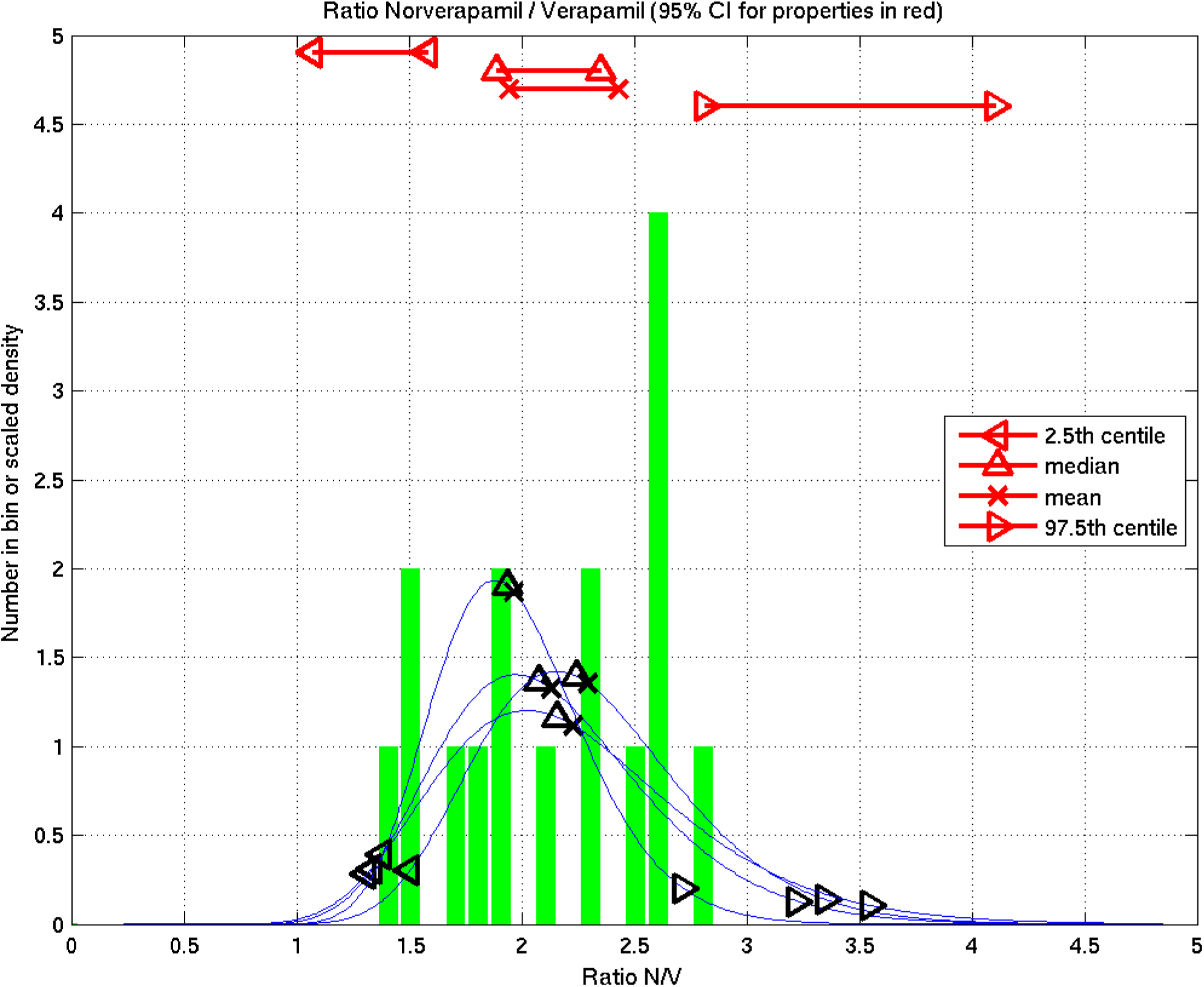
Inference from the present study on the ratio of norverapamil AUC to verapamil AUC: The green histogram represents the individual values for each patient. Each blue curve represents a sample from the posterior distribution given that data of the distribution in the population. For each blue curve, the black left-pointing triangle is the 2.5th centile, the up-pointing triangle the median, the cross the mean, and the right-pointing triangle the 97.5th centile. In red at the top of the figure are the central 95% Bayesian confidence intervals for each of these quantities, based on 9000 samples of the distribution over the population (i.e. on 9000 such blue curves).

## 7 The fifth problem – comparing rifampicin or isoniazid levels before and after verapamil

As above, we made the assumption that these were log-Gaussianly distributed.

For this, as in section 5, we used the conjugate joint prior to (*µ, S*), the mean and scale (inverse of covariance) parameters of a 2-d Gaussian, given by

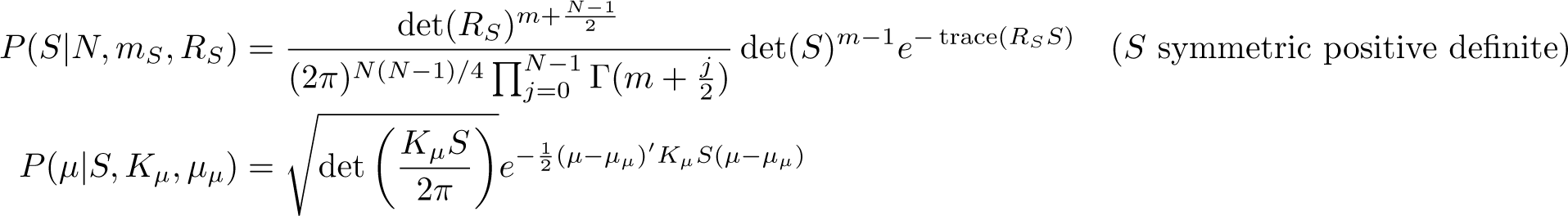

We set the broad spherically symmetric prior (suitably scaled for this data in logarithms of quantities in mg hours / litre) given by:

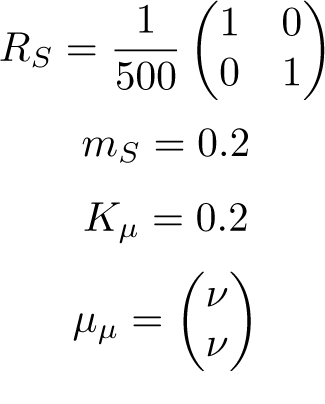

where for analysing the data on rifampicin (resp. isoniazid) set *ν* to be the mean of the logs of all the rifampicin (resp. isoniazid) AUCs (both pre and post verapamil).

We then considered the data composed of the logarithms of raw AUC values for rifampicin (resp. isoniazid) before and after verapamil. Since we had 10 patients contributing, this gave us a 2 *×* 10 array, which we treated as ten 2-d vectors assumed to be from a Gaussian. We calculated the posterior distribution of the parameters of that Gaussian using Bayes’ theorem, and drew 10,000 samples of the resulting 2-d samples of *µ*. We reported the fraction of samples whose second coordinate was greater than its first as the posterior probability that the geometric mean post-verapamil was greater than that pre-verapamil. We reported the relevant exponentiated centiles of the first coordinate of the samples for the pre-verapamil, and of the second coordinate of the samples for the post-verapamil, as the confidence limits of the geometric mean.

## Statistical Appendix 2 Statistical Definitions and Abbreviations

Version 1.12

### 1 Definitions and abbreviations

We use the following definitions, including some which make abbreviated phraseology precise that would otherwise be ambiguous.

1. **Posterior probability** – the probability given the data, i.e. after seeing the data, or in the light of the data.

2. **Restricted geometric mean** – the geometric mean of a distribution, restricted to an interval excluding only extreme tail events; see Appendix 2 of [1] for reasons for using this concept.

3. **A is significantly greater than B (** ≥ *α*) – the posterior probability (see 1) that the geometric mean of the variable A in the relevant population was greater than that of the variable B was at least *α*, where *α ≥* 0.95 (and similarly for “significant increase”, etc.).

4. **A is significantly greater than each B in list** (*≥ α*) – for each B in list, A is significantly greater than B (*≥ α*) (see 3).

5. **A is not significantly different from B (** ≤ *α*) – the posterior probability (see 1) that the geometric mean of the variable A in the relevant population was greater than that of the variable B was greater than 1 − *α* and below *α*, where 0.5 ≤ *α <* 0.95 (and similarly for “no significant difference”, etc.).

6. **A is not significantly different from each B in list** (*≤ α*) – for each B in list, A is not significantly different from B (*≤ α*) (see 5).

7. **The probability that A was greater than B was *p*** – the posterior probability (see 1) that the geometric mean of the variable A in the relevant population was greater than that of the variable B was *p*.

8. **The probability that A was greater than each B in list was** ≥ *p* – for each B in list, the posterior probability (see 1) that A was greater than B (see 7) was at least *p*. In particular this does *not* mean “the posterior probability was at least *p* that, for all B in list, A was greater than B”, as this probability is usually very much smaller.

9. **A was greater than B** (where no probability or significance is mentioned) – the geometric mean of the data for A was greater than that for B.

10. **Probability value** – In frequentist statistics, the “p-value” of an analysis is given as the probability of the observed data falling (as it did) into a predefined critical region if the null hypothesis is true (or sometimes one minus this probability).

In a Bayesian analysis the posterior probability of something is the probability that that that something is true given the data.

In order to avoid confusion, we avoid giving such Bayesian probabilities in the form e.g. “(*p*=0.97)”, and simply state the probability of something being true as e.g. “(0.97)”.

Thus if the probability that A is greater than B is 0.02, then the probability that B is greater than A is 0.98, so B is significantly greater than A and A is significantly less than B (see 3).

There are no frequentist “p-values” given in this paper.

11. **Reporting format** We report inferred values of an unknown quantity X in the form (*a*, *b* — *c*), where *a* is the posterior arithmetic mean of the geometric mean of X over the population, *b* is the 2.5th posterior centile of the geometric mean of X over the population, and *c* is the 97.5th posterior centile of the geometric mean of X over the population. Then (*b* — *c*) is the equitailed 95% Bayesian confidence interval for the geometric mean of X over the population. We do similarly for plotted values showing a central estimate and a confidence interval.

To clarify this for those not used to Bayesian analyses, we refer to Figure 4 of Appendix 1 of this paper, and proceed as follows. We believe that X is distributed over the population according to a log-Gaussian (a.k.a. log-normal) distribution

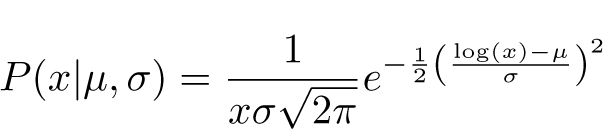

where the parameters *µ* and *σ* are unknown. This distribution has (arithmetic) mean *e^µ^*^+^ ^1 *σ*2^, mode *e^µ−σ^*^2^, and geometric mean and median both given by *e^µ^*.

Now, we would like to be able to report the value of *g* = *e^µ^*, the geometric mean of this distribution over the population, but we do not know it. So, as Bayesians, we calculate the posterior distribution of *g*, by drawing thousands of samples from the posterior distribution on (*µ, σ*) given the data (using the methods described in appendix 1 section 6). For each such sample we can plot one of the blue curves in the figure, and for each such sample we can calculate *g* = *e^µ^*, giving a corresponding sample of *g*. The overall set of such samples then allows us to calculate the arithmetic mean and centiles of the posterior distribution of *g*.

In other words we do not know which of the blue curves in the figure (or many others like them) is the true distribution of X over the population. Each of these curves has its geometric mean (and median) marked by an upward pointing black triangle. By considering thousands of sample blue curves from the posterior distribution on the distribution of X, we get thousands of sample values of the geometric mean *g* over the population, from which we can deduce the posterior arithmetic mean and centiles of *g*. The upward pointing red triangles at the top of the figure then show the 2.5th and 97.5th centiles of g.

12. **The restricted geometric mean of A was significantly greater than that of B** (≥α): – the posterior probability (see 1) that the restricted geometric mean (see 2) of A was greater than that of B was at least α.

13. **The restricted geometric mean of A was significantly greater than those of each B in list (≥ α): – for each B in list**, the restricted geometric mean of A was significantly greater than that of B (≥ α) (see 12, and note also the “in particular” part of 8).

